# Genotype-stratified GWAS meta-analysis reveals novel loci associated with alcohol consumption

**DOI:** 10.1101/2021.06.02.21258094

**Authors:** Yuriko N. Koyanagi, Masahiro Nakatochi, Hidemi Ito, Yumiko Kasugai, Akira Narita, Takahisa Kawaguchi, Hiroaki Ikezaki, Asahi Hishida, Megumi Hara, Toshiro Takezaki, Teruhide Koyama, Haruo Mikami, Sadao Suzuki, Sakurako Katsuura-Kamano, Kiyonori Kuriki, Yasuyuki Nakamura, Kenji Takeuchi, Atsushi Hozawa, Kengo Kinoshita, Yoichi Sutoh, Kozo Tanno, Atsushi Shimizu, Isao Oze, Yukino Kawakatsu, Yukari Taniyama, Issei Imoto, Yasuharu Tabara, Meiko Takahashi, Kazuya Setoh, Shiori Suzuki, Atsushi Goto, Ryoko Katagiri, Taiki Yamaji, Norie Sawada, Shoichiro Tsugane, Kenji Wakai, Masayuki Yamamoto, Makoto Sasaki, Fumihiko Matsuda, Motoki Iwasaki, Paul Brennan, Keitaro Matsuo

## Abstract

An East Asian-specific variant on *aldehyde dehydrogenase 2* (*ALDH2* rs671, G>A) is the major genetic determinant of alcohol consumption. We performed an rs671 genotype-stratified genome-wide association study meta-analysis in up to 40,679 individuals from Japanese populations to uncover additional loci associated with alcohol consumption in an rs671-dependent manner. No loci satisfied the genome-wide significance threshold in wild-type homozygotes (GG), but six loci (*ADH1B, ALDH1B1, ALDH1A1, ALDH2, GOT2*, and *MYOM1-MYL12A*) did so in heterozygotes (GA). Of these, three loci (*ALDH2, GOT2*, and *MYOM1-MYL12A*) were novel, and two (*ADH1B* and *ALDH1B1*) showed genome-wide significant interaction with rs671. Our results identify a new genetic architecture associated with alcohol consumption, and shed additional light on the genetic characteristics of alcohol consumption among East Asians.

---

Alcohol consumption is a major contributor to mortality and influences risk for various human diseases and disorders^1^. Even moderate consumption may have a substantial impact on mortality^2^. Indeed, the latest Global Burden of Disease study on alcohol use states that the level of consumption should be reduced to zero to minimize health risk^1^. Alcohol consumption has been considered a heritable trait^3,4^. The number of genetic studies on alcohol consumption is increasing^5-14^, and the genetic variants that are consistently reproducible are those of genes encoding alcohol-metabolizing enzymes^5,7,8,10-14^. Ingested alcohol is predominantly metabolized to acetaldehyde through alcohol dehydrogenase (ADH) enzymes, and aldehyde dehydrogenase (ALDH) enzymes further catalyze the oxidation of acetaldehyde to acetate^15^. Notably, rs671 (c.1510G>A [p.Glu504Lys]), a functional single nucleotide polymorphism (SNP) in the *ALDH2* gene which is highly prevalent in East Asians^16^, is a strong and well-known determinant of alcohol consumption. Every previous genome-wide association study (GWAS) in East Asians^5,7,8,11,14^ identified the strongest signals in the rs671 variant (or variants in strong linkage disequilibrium [LD] with rs671), ranging from *P* <1.0 × 10^−58^ (*n* = 2,834)^5^ to *P* <1.0 × 10^-4,740^ (*n* = 165,084)^14^.

Among ALDH isoforms, ALDH2 has by far the highest affinity for acetaldehyde (Km <1 µM) and is primarily responsible for its oxidation^16,17^. Because the *ALDH2* rs671variant inactivates ALDH2 enzymatic activity, individuals who are heterozygous (GA) or homozygous (AA) for this variant experience a rapid accumulation of blood acetaldehyde after alcohol ingestion^16^. This variant thereby increases exposure to the unpleasant effects of acetaldehyde (e.g. flushing, headache, palpitation, and nausea), which in consequence significantly reduces alcohol consumption and thereby confers a protective effect against alcohol-induced carcinogenesis^18^. Conversely, heterozygotes (GA) who drink alcohol experience increased susceptibility to carcinogenesis, in particular for head and neck and esophageal cancers, due to higher concentrations of acetaldehyde, one of the most likely carcinogens in alcohol^15^. With regard to variant homozygotes (AA), however, these have rarely evidenced an increased cancer risk associated with alcohol, because they are unable to oxidize acetaldehyde, a characteristic which is highly correlated with nondrinking^19,20^. In contrast, heterozygotes having 16-18% of normal enzyme activity^21,22^ show a broader range of alcohol consumption. Overall, the highest risk group for alcohol-related cancers are heterozygotes^23,24^, and alcohol consumption among genotypes of rs671 shows distinct genetic heterogeneity.

Here, to further decipher the genetic architecture of alcohol consumption in consideration of the status of this unique variant of rs671, we conducted a meta-analysis of rs671 genotype-stratified GWASs comprising up to 40,679 Japanese individuals. Using rs671 genotype-stratified analyses, we tested the hypothesis that variants associated with alcohol consumption exhibit rs671 genotype-dependent associations, and sought novel variants conferred by genetic interaction of the rs671 genotype. We considered that this approach might help uncover loci with differential influence on alcohol consumption among genotypes, and enable the detection of loci whose effects were indistinct in previous GWASs.

## Results

### Characteristics of study participants

A total of 40,679 individuals were included in this GWAS meta-analysis of five Japanese cohorts, namely the Hospital-based Epidemiologic Research Program at Aichi Cancer Center (HERPACC)^25^ Study (*n* = 4,958), the Japan Multi-Institutional Collaborative Cohort (J-MICC)^26,27^ Study (*n* = 13,236), the Japan Public Health Center-based Prospective (JPHC)^28^ Study (*n* = 10,037), the Tohoku Medical Megabank Community-Based Cohort (TMM)^29^ Study (*n* = 7,857), and the Nagahama Prospective Cohort for Comprehensive Human Bioscience (Nagahama) Study^30^ (*n* = 4,591), after imputation and quality control of individual subject genotype data (Supplementary Information and Supplementary Tables 1 and 2). Median self-reported daily alcohol intake, mean age of participants, number of ever/never drinkers, and proportion of male participants were obtained from the studies and are shown in Supplementary Table 1. The number of participants included in each analysis for daily alcohol intake and drinking status was as follows: rs671 wild-type homozygotes (GG)-only analysis, *n* = 23,398 and 24,514; heterozygotes (GA)-only analysis, *n* = 13,385 and 13,848; unstratified analysis, *n* = 39,077 and 40,679; and interaction analysis, *n* = 36,783 and 38,362, respectively (Supplementary Table 3). As the number of variant homozygotes (AA) was small (*n* = 2,294 for daily alcohol intake; *n* = 2,317 for drinking status) and included only 101 subjects in the ever drinking group (Supplementary Table 3), association analysis in variant homozygotes only was not conducted, and these subjects were excluded from the interaction analyses. Quantile-quantile plots revealed no evidence of genomic inflation (Supplementary Figures 1 and 2) and the genomic inflation factors ranged from 0.99 to 1.02 for meta-analyses.

### *ALDH2* rs671 genotype-stratified, unstratified, and interaction GWAS meta-analyses

Major results of the genotype-stratified GWAS meta-analysis are summarized in Figures 1-4. In wild-type homozygotes (GG), no genome-wide significant loci were detected for either daily alcohol intake or drinking status (Figures 1a and 2a). In heterozygotes (GA), on the other hand, six and four loci satisfied the genome-wide significance (*P* < 5.0 × 10^−8^) for daily alcohol intake and drinking status, respectively (Figures 1b, 2b, 3 and 4). These included three loci that were previously implicated in alcohol consumption, namely chromosome 4q23^10-14^, *ALDH1B1*^14^, and *ALDH1A1*^14^. Of the three remaining loci, two loci (*GOT2* at 16q21, and *MYOM1-MYL12A* at 18p11.31) have not been reported in previous GWASs of alcohol consumption, and one locus (chromosome 12q24.12) is the same locus as rs671. Regional association plots for these novel loci are shown in Figure 5. The lead SNPs are rs56884502 in chromosome 12q24.12 (*ALDH2*), rs73550818 in *GOT2*, and rs572435541 in *MYOM1-MYL12A* for daily alcohol intake, and rs7978737 in chromosome 12q24.12 (*ACAD10-ALDH2*) for drinking status. The unstratified GWAS showed three hits (rs1260326 in *GCKR* for daily alcohol intake; rs1229984 in *ADH1B* and rs671 in *ALDH2* for daily alcohol intake and drinking status) (Figures 1c, 2c, 3 and 4), all of which have been previously reported^5,7-14^. Regional association plots for all identified regions other than those in Figure 5 are shown in Supplementary Figures 3 and 4.

**Figure 1.**
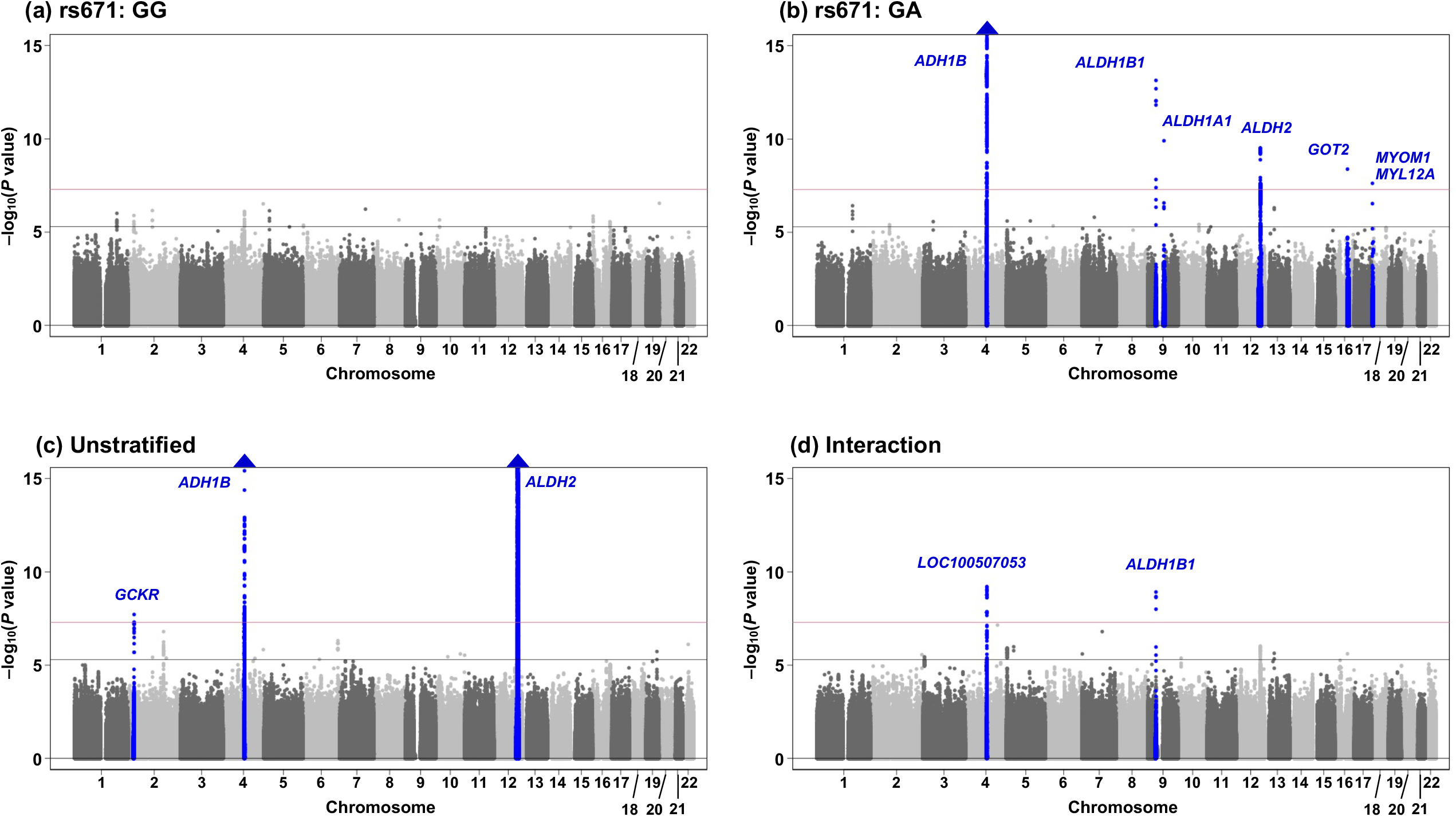
Manhattan plots of the GWAS of daily alcohol intake. The results for (a) rs671 wild-type homozygotes (GG), (b) rs671 heterozygotes (GA), (c) unstratified, and (d) interaction with rs671 are shown. The position on each chromosome (x-axis) and the observed –log_10_(*P* value) (y-axis) of all tested genetic variants are shown. The solid red and gray lines indicate genome-wide and suggestive significance levels, respectively. Blue triangles represent loci containing SNPs with *P* values of <1×10^−15^.

**Figure 2.**
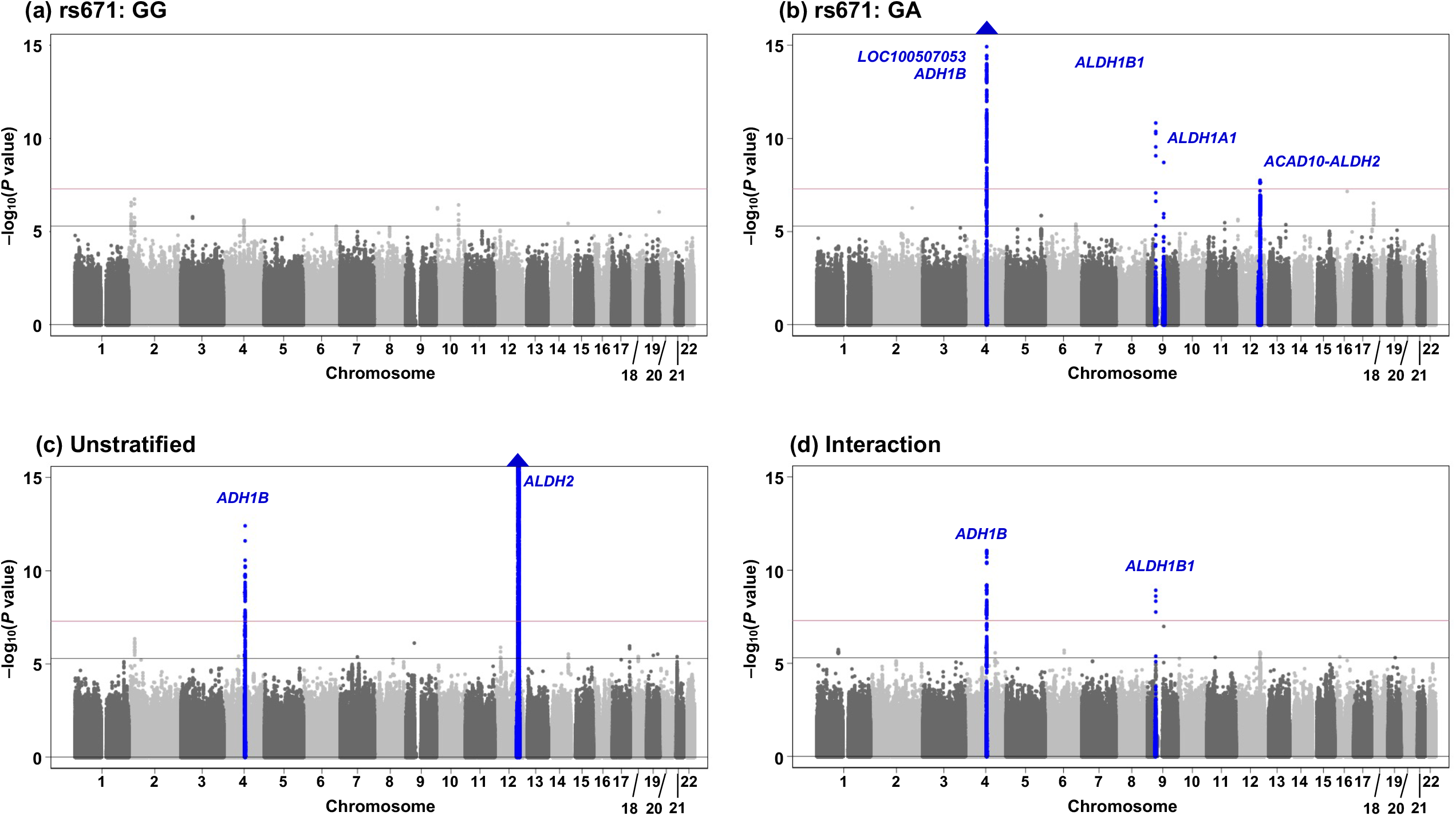
Manhattan plots of the GWAS of drinking status. The results for (a) rs671 wild-type homozygotes (GG), (b) rs671 heterozygotes (GA), (c) unstratified, and (d) interaction with rs671 are shown. The position on each chromosome (x-axis) and the observed –log_10_(*P* value) (y-axis) of all tested genetic variants are shown. The solid red and gray lines indicate genome-wide and suggestive significance level, respectively. Blue triangles represent loci containing SNPs with *P* values of <1×10^−15^.

**Figure 3.**
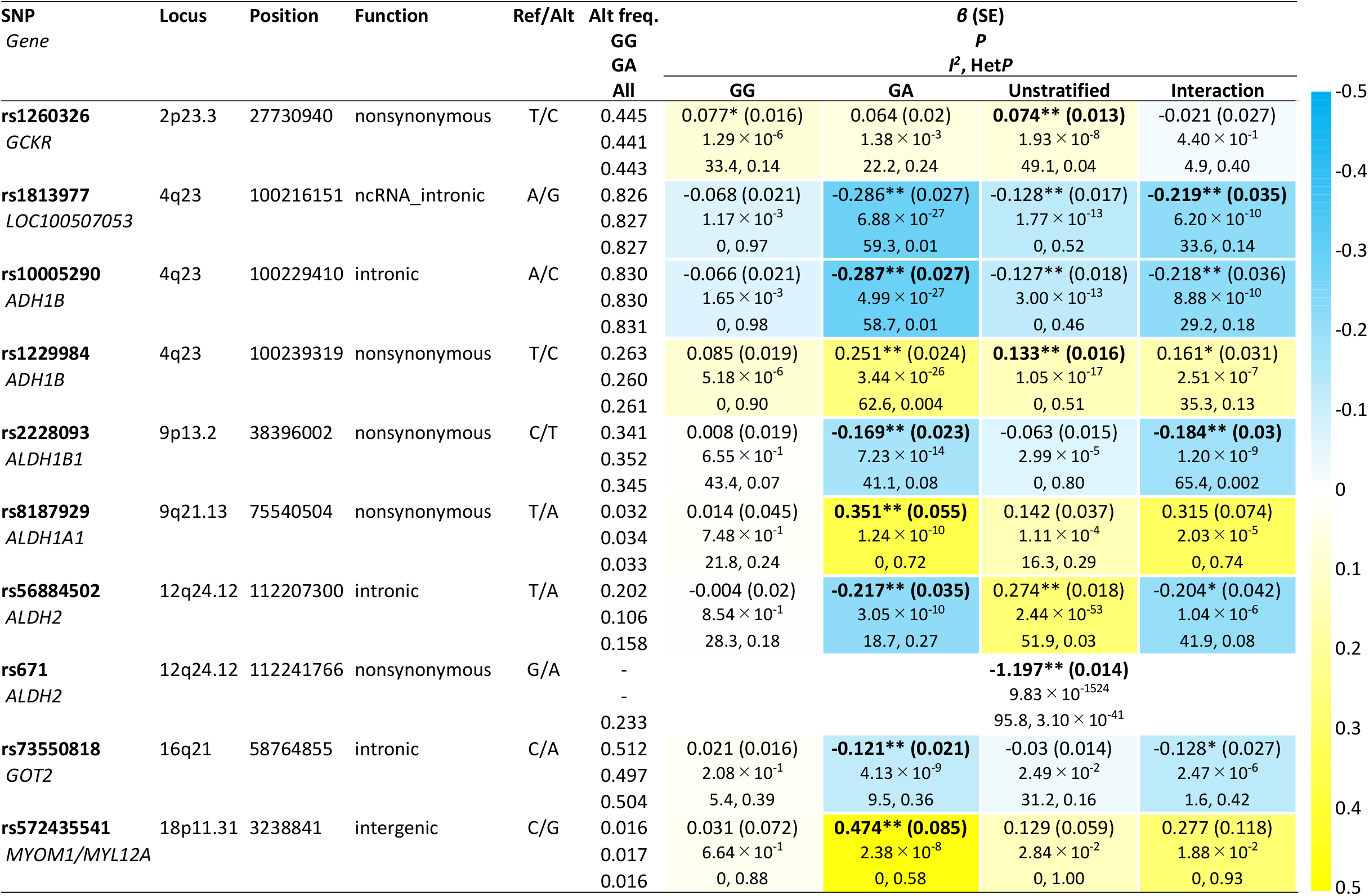
Genomic loci reaching genome-wide significance in either analysis for association with daily alcohol intake. Direction of effects of identified variants other than rs671 is presented as a heatmap. Estimates with a single asterisk show suggestive significance (*P* < 5.0 × 10^−6^). Estimates with double asterisks show genome-wide significance (*P* < 5.0 × 10^−8^). Lead SNP in each locus is highlighted with its estimates in bold. SNP, single nucleotide polymorphism; Ref, reference allele; Alt, alternative allele; freq., frequency; SE, standard error; Het*P, P* value from test of heterogeneity.

**Figure 4.**
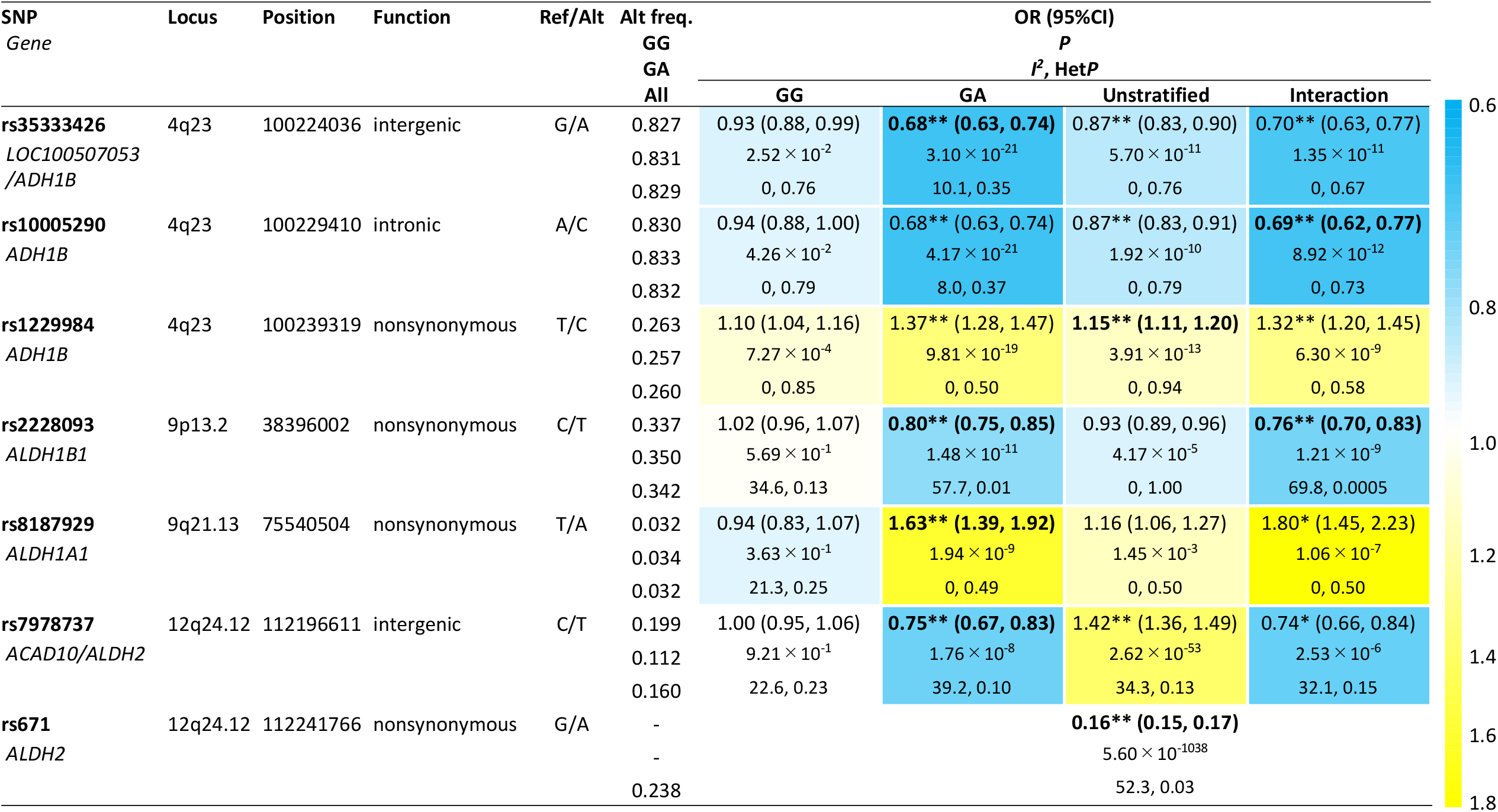
Genomic loci reaching genome-wide significance in either analysis for association with drinking status. Direction of effects of identified variants other than rs671 is presented as a heatmap (with colors indicating associated normalized ORs). Estimates with a single asterisk show suggestive significance (*P* < 5.0 × 10^−6^). Estimates with double asterisks show genome-wide significance (*P* < 5.0 × 10^−8^). Lead SNP in each locus is highlighted with its estimates in bold. SNP, single nucleotide polymorphism; Ref, reference allele; Alt, alternative allele; freq., frequency; OR, odds ratio; 95% CI, 95% confidence interval; Het*P, P* value from test of heterogeneity.

**Figure 5.**
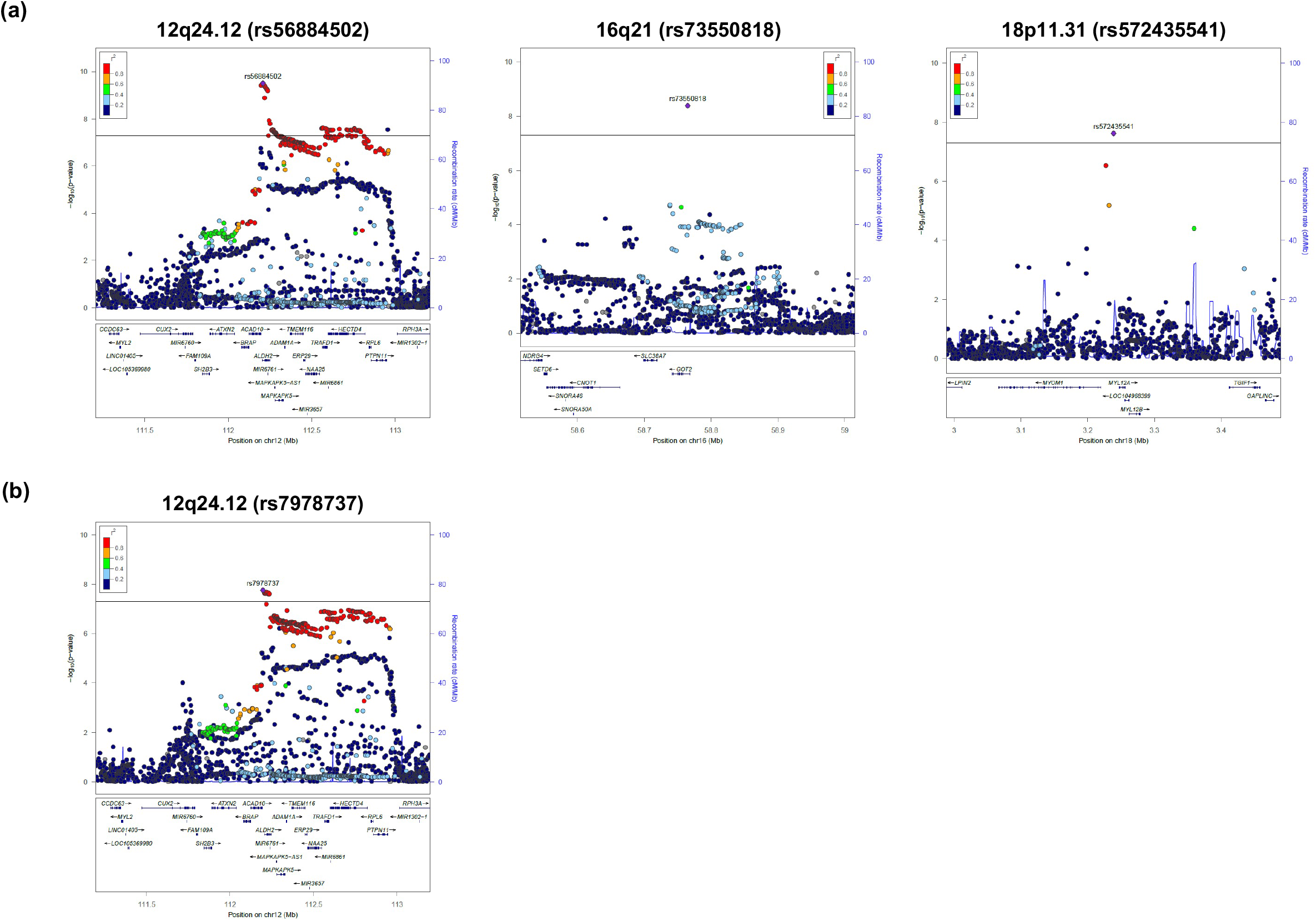
Regional association plots of the identified novel regions. Regional association plots for (a) daily alcohol intake and (b) drinking status in rs671 heterozygotes are shown. The vertical axis indicates the –log_10_(*P* value) for the assessment of the association of each SNP with daily alcohol intake or drinking status. The black line represents a genome-wide significance threshold of 5.0×10^−8^. The colors indicate the LD (*r*^*2*^) between each lead SNP and neighboring SNPs based on the JPT population in the 1000 Genomes Project Phase 3.

A further analysis evaluating variant-rs671 interaction detected two loci (chromosome 4q23 and *ALDH1B1*) showing genome-wide significant interaction with rs671 (Figures 1d, 2d, 3 and 4 and Supplementary Figures 3c and 4c). This is the first identification of the interactive effects of these two loci on rs671, although that of rs1229984 in *ADH1B* at 4q23 on rs671 was suggested from a meta-analysis of studies using candidate gene-based approaches^31^. Among the other loci reaching genome-wide significance in either rs671-stratified or unstratified analysis, three loci (*ALDH1A1*, chromosome 12q24.12, and *GOT2*) demonstrated interaction with rs671 with a suggestive significance level (*P* < 5.0 × 10^−6^) for daily alcohol intake and/or drinking status (Figures 3 and 4). Regarding the multiple hits on chromosome 4q23 observed through these analyses, rs1813977 (interaction), rs35333426 (for drinking status among heterozygotes), and rs10005290 (for daily alcohol intake among heterozygotes) were in strong LD with a functional SNP of *ADH1B* (rs1229984) (all *r*^*2*^ and *D’* values are 0.71 and 1.00 in 1000 Genome Project phase 3-Japanese in Tokyo (1KGP-JPT), respectively). We further applied a random effects model^32^ given that estimates in *ALDH1B1* rs2228093 showed between-study heterogeneity (*P* values from test of heterogeneity <0.05) (Figures 3 and 4), but the results did not change substantially (*P* = 8.34 ×10^−11^ in the interaction analysis for daily alcohol intake; and *P* = 4.38 ×10^−12^ and *P* = 1.62 ×10^−11^ in the analyses of heterozygotes only and interaction, respectively, for drinking status).

Supplementary Table 4 shows functional annotation results and allele frequencies across different ancestries for the lead SNPs. Five SNPs, namely rs1260326 on *GCKR*, rs1229984 on *ADH1B*, rs2228093 on *ALDH1B1*, rs818787929 on *ALDH1A1*, and rs671 on *ALDH2*, are non-synonymous. According to the 1KGP database, three SNPs (rs8187929, rs671, and rs572435541) are polymorphic, with a minor allele frequency (MAF) of 0.046, 0.170, and 0.009 in the East Asian (EAS) population, respectively. In contrast, they are monomorphic in the European (EUR) population. The *ADH1B* rs1229884 C allele is major in the EUR population (AF = 0.970) but minor in the EAS population (AF = 0.300).

### Effect of a novel SNP within the same locus as rs671

Figures 3 and 4 show the direction of effects of the identified variants other than rs671 under each analysis. Notably, with regard to the novel SNP in chromosome 12q24.12 (*ALDH2* rs56884502, T>A), the A allele of rs56884502, which was associated with decreasing daily alcohol intake in the rs671 heterozygotes (*β* = −0.217), showed the opposite direction of effect in the unstratified analysis (*β* = 0.274) (Figure 3). This apparently conflicting result was due to strong LD between rs56884502 and rs671. The 1KGP-JPT (*n* = 104) and our own direct genotyped data from subjects in the HERPACC Study (*n* = 96) indicated that there were only three rs56884502-rs671 haplotypes, namely T-G, A-G and T-A (Supplementary Table 5). The respective LD coefficients of *r*^*2*^ and *D’* were <0.1 and 1.0 (Supplementary Table 5). Results from LD analysis based on the 1KGP JPT data (*n* = 104) of rs671 and the 73 SNPs at 12q24.12 which showed genome-wide significance for daily alcohol intake among the heterozygotes (Supplementary Table 6) are shown in Supplementary Figure 5. The pairwise *D’* figure showed that all 73 SNPs were in complete LD in terms of *D’* (Supplementary Figure 5). Further, the three indicated haplotypes of rs671 and these SNPs could explain >99% (Supplementary Figure 6). Therefore, when evaluated without stratification, rs56884502 A allele, which formed a haplotype with rs671 G allele only, was associated with increasing drinking intensity, by reflecting the effect of rs671 G allele. However, when stratified, rs56884502 A allele turned out to have the opposite direction of effect–decreasing drinking intensity. The other lead SNP in chromosome 12q24.12 for drinking status (rs7978737) was in LD with rs56884502 (*r*^*2*^ = 0.97 in 1KGP JPT), and accordingly showed the same phenomenon (Figure 4).

### Associations of previously reported loci

Among the previously reported loci in the EUR population^9,11,12,33,34^ other than those replicated with a genome-wide significance level in this study, we observed nominal evidence of association (*P* < 0.05) for nine loci in the unstratified analysis, six loci in wild-type homozygotes, and seven loci in heterozygotes (Supplementary Table 7).

### eQTL analysis of novel SNPs

Of the detected novel variants, rs56884502 and rs7978737 in chromosome 12q24.12 and rs73550818 in *GOT2* were found to be eQTL using the GTEx database (Supplementary Table 8). rs56884502 A allele and rs7978737 T allele are associated with decreased expression of *ALDH2* in multiple tissues. rs73550818 A allele is associated with increased expression of *GOT2* in liver (*P* = 1.0×10^−8^).

## Discussion

We report here the results of an rs671 (G>A) genotype-stratified GWAS meta-analysis of alcohol consumption with a total of 40,679 participants from five Japanese cohorts. While three loci (*GCKR, ADH1B*, and *ALDH2*) were identified in the unstratified analysis, the rs671 genotype-stratified GWAS identified no loci in wild-type homozygotes (GG) and six loci (*ADH1B, ALDH1B1, ALDH1A1, ALDH2, GOT2*, and *MYOM1-MYL12A*) in heterozygotes (GA). Of these, three loci (*ALDH2* at 12q24.12, *GOT2* at 16q21, and *MYOM1-MYL12A* at 18p11.31) are novel in the context of alcohol consumption. Further, the interaction GWAS identified for the first time two loci (*ADH1B* and *ALDH1B1*) showing genome-wide significant interaction with rs671.

The failure of other loci to reach a genome-wide significant level in rs671 wild-type homozygotes indicates that the rs671 GG genotype itself is strong enough to make a significant contribution to determining the alcohol consumption phenotype in this population. This is consistent with the observation that this phenotype has been resistant to gene discovery efforts in non-Asian populations, where rs671 is often monomorphic^35^. Further, the strongest signal for daily alcohol intake in rs671 heterozygotes was observed in *ADH1B* (*P* < 5.0 × 10^−26^), followed by *ALDH1B1* (*P* = 7.2 × 10^−14^) and then *ALDH1A1* (*P* = 1.2 × 10^−10^), all of which are associated with the concentration of acetaldehyde. ADH1B is the predominant isoform involved in alcohol oxidation, whereas ALDH1B1 and ALDH1A1 are the ALDH isoforms involved in acetaldehyde oxidation, with the second (Km 30 µM) and third (Km 50–180 µM) highest affinities for acetaldehyde, respectively^17^.

The nonsynonymous lead SNP of rs1229984 (T>C [p.His48Arg]) found in the *ADH1B* coding region is associated with slow alcohol metabolism, leading to the slow accumulation of acetaldehyde and consequently greater alcohol consumption^19,20^. Although rs2228093 (C>T [p.Ala86Val]) in *ALDH1B1* and rs8187929 (T>A [p.Ile177Phe]) in *ALDH1A1* were first shown to be associated with drinking status in a previous Japanese GWAS^14^, their effects on enzyme activity are not fully understood. However, rs2228093 in *ALDH1B1* was also shown to possibly influence alcohol consumption in European populations using a candidate gene approach^36,37^. In addition, the protective effect of rs2228093 T allele against alcohol consumption observed in this study is consistent with the results of previous studies using bioinformatic analyses, which predicted disruption of the structural flexibility of the protein product^38^ and catalytic inactivity^39^ of ALDH1B1 in the presence of the rs2228093 T allele. Our present study genetically confirmed that, at least in this population, alcohol consumption level is largely determined by the concentration of acetaldehyde, because no significant signal was detected in rs671wild-type homozygotes whereas signals in the genes encoding the second and third enzymes involved in the concentration of acetaldehyde were identified in heterozygotes. Elucidating the functional contributions of rs2228093 in *ALDH1B1* and rs8187929 in *ALDH1A1* to alcohol/aldehyde metabolism requires further investigation.

Using a stratified method based on rs671 genotype, we were able to uncover the effect of a novel SNP at the same locus as rs671. ALDH2 is a tetramer which is regarded as a dimer of dimers. The rs671 A allele is predicted to disrupt the structure of not only its own subunit but also its dimer partner, reducing the stability of the tetramic structure of ALDH2 and resulting in a dramatic reduction in enzyme activity^40^. This East Asian-specific SNP is considered to be a relatively young polymorphism^41^ and to have been under strong recent natural selection pressure in the Japanese population^42^. On the other hand, the novel SNP at 12q24.12 of rs56884502 is a globally common SNP located <40Kb distant to rs671 (Supplementary Table 4). Given this evidence and the two LD measures of *r*^*2*^ <0.1 and *D’* = 1.0 for rs56884502 and rs671, we speculate that rs56884502 arose prior to rs671, and that rs671 then arose on a different branch from rs56884502 in the rs56884502-rs671 T-G haplotype without subsequent historic recombination, finally resulting in the three haplotypes of T-G, A-G, and T-A. The protective effect of the rs56884502 A allele against alcohol consumption observed in rs671 heterozygotes can therefore be regarded as a protective effect of the rs56884502-rs671 T-A/A-G diplotype. Accordingly, we hypothesize that rs56884502 (or variants in LD) is associated with reduced enzyme activity via an effect on the rs671 A allele located on the opposite haplotype. However, rs56884502 is located in the intron of *ALDH2* (Supplementary Table 4) and the rs56884502 A allele was found to be associated with decreased expression of *ALDH2* (Supplementary Table 8). Further, none of the other 72 SNPs that were in LD with rs56884502 and showed genome-wide significance for daily alcohol intake are located within the coding region (Supplementary Table 6), suggesting that while these SNPs may be potentially associated with expression, they may have no direct effect on the protein structure or tetramer formation of ALDH2 by interacting with the rs671 A allele on the opposite haplotype. Further, this potential effect on *ALDH2* expression is inconsistent with the lack of protective effect of the rs56884502 A allele in a population with rs671 wild-type homozygosity. One possible explanation is that this protective effect may be too small for detection in wild-type homozygotes, but if the causative exonic SNP may be hidden and the variant is rare, this would be difficult or impossible to impute using the 1KGP reference panel. Further elucidation of this rs671-dependent protective effect of rs56884502 will require deep whole-genome sequencing-based analysis and/or experimental study.

Our genome-wide analysis indicates the interactive effect of rs2228093 in *ALDH1B1* with rs671. ALDH1B1 is another mitochondrial ALDH which shares 75% similarity with ALDH2 at the amino acid sequence level, and is predicted to form a homotetramer, similarly to ALDH2^43^. Although ALDH1B1 is also able to oxidize acetaldehyde, individuals with the rs671 A allele are reported not to exhibit a compensatory increase in ALDH1B1 activity^44^. Further, a bioinformatics analysis predicted protein-protein interactions between ALDH2 and ALDH1B1, indicating that ALDH2 and ALDH1B1 subunits are likely to form heterotetramers^44^. These findings suggest the hypothesis that the rs671 A allele reduces the catalytic activity of ALDH1B1. They also explain the present finding of gene-gene interaction between the rs671 A allele and rs2228093 T allele, both of which are validated and predicted^39^ to be associated with catalytic inactivity. Moreover, these findings may further explain the limited and conflicting genetic evidence for rs2228093 on drinking in European populations^36,37^, in which rs2228093 is polymorphic (MAF = 0.15 in 1KGP EUR) but rs671 is monomorphic.

The remaining newly identified loci associated with alcohol consumption in this study are *GOT2* and *MYOM1-MYL12A*. The lead SNPs in these loci are located within the non-coding region and the functional effects of these variants are unknown. However, the lead SNP in *GOT2* (rs73550818, C>A) is located in the intron of *GOT2* and showed a protective effect against alcohol consumption in heterozygotes; this may be a suitable target for future study, given that the rs73550818 A allele was shown to be significantly associated with increased levels of aspartate aminotransferase (AST), a biochemical marker for liver injury, in a previous GWAS of 134,154 Japanese individuals^45^. *GOT2* encodes the mitochondrial isoform of glutamic-oxaloacetic transaminase; this plays an important role in many processes, including amino acid metabolism, long-chain fatty acid uptake, and the urea and tricarboxylic acid cycles^46^. An *in vivo* study suggested that increased mitochondrial AST among alcoholics is a consequence of the pharmacologic upregulation of *GOT2* gene expression by ethanol, which further mediates fatty acid uptake, resulting in alcoholic fatty liver^46^. Considering that the rs73550818 A allele is associated with increased expression of *GOT2* in the liver (Supplementary Table 8), this allele might be associated with ethanol-induced liver injury. On the other hand, previous studies of rs671 showed significantly lower AST in heterozygotes than in wild-type homozygotes among drinkers^47^, even after adjustment for alcohol intake^7^. An observational study of patients with alcoholic liver injury^48^ and a study of *Aldh2* knockout mice^49^ suggested a protective effect of the rs671 A allele on ethanol-induced liver injury. These findings suggest the opposite effects of the rs73550818 A and rs671 A alleles on ethanol-induced liver injury. The mechanism of the suggested interaction between rs73550818 and rs671 observed in our present study therefore warrants further investigation.

This study has several strengths. First, most of the included cohorts were population-based and included a large number of general Japanese individuals, and the possibility of selection bias is likely small. Second, the study involved a single ethnic group with a similar religious and cultural background, making it unlikely that these factors would bias the phenotype of alcohol consumption^50^. An important limitation is that data on alcohol consumption were self-reported. Nevertheless, these data were collected at baseline survey using validated questionnaires or their variants in all studies. Any misclassification bias is therefore likely to be non-differential, in which case the validity of our observed associations is likely to hold.

Finally, we would like to note a merit of this particular type of genotype-stratified GWAS. If there is a phenotype of interest and a polymorphism that has a strong influence on that phenotype – in this case, alcohol consumption is the phenotype on which *ALDH2* rs671 has a decisive effect - this method is highly effective. The fact that many of the polymorphisms revealed in this study are related to alcohol metabolism may strongly support this notion. Although GWAS was originally conceived as hypothesis-free, the hypothesis-driven approach we used here worked effectively, indicating its potential in the search for new targetable loci. A phenomenon observed in this study is generally termed gene-gene interaction or SNP-SNP interaction, and its existence has been identified using the candidate approach^51^. GWASs examining interactions with environmental factors using a statistical interaction term are not necessarily successful: in this study, the interaction term approach was not effective despite use of a strong partner, rs671. Accordingly, we propose that hypothesis-based genotype-stratified GWAS represents a promising new approach to discovery.

In conclusion, we performed an *ALDH2* rs671 genotype-stratified GWAS and successfully identified several loci that were associated with alcohol consumption in an rs671-dependent manner. This study further reveals the genetic structure of alcohol consumption, and should deepen our knowledge of the pathogenesis of alcohol-related diseases and disorders.

## Methods

### Study subjects and genotyping

We performed a genome-wide meta-analysis based on the Japanese Consortium of Genetic Epidemiology studies (J-CGE)^52^ and the Nagahama Study^30^, both of which comprised general Japanese populations. The J-CGE consisted of the following Japanese population-based and hospital-based studies: the HERPACC Study^25^, the J-MICC Study^26,27^, the JPHC Study^28^, and the TMM Study^29^. Individual study descriptions and an overview of the characteristics of the study populations are provided in the Supplementary Information and Supplementary Table 1. Data and sample collection for the participating cohorts were approved by the respective research ethics committees. All participating studies obtained informed consent from all participants by following the protocols approved by their institutional ethical committees.

### Phenotype

Information on alcohol consumption was collected by questionnaire in each study. Because the questionnaires were not homogeneous across the studies, we harmonized the two alcohol consumption phenotypes of drinking status (never versus ever drinker) and daily alcohol intake (g/day) in accordance with each study’s criterion. Details are provided in the Supplementary Information.

### Quality control and genotype imputation

Quality control for samples and SNPs was performed based on study-specific criteria (Supplemental Table 2). Genotype data in each study were imputed separately based on the 1000 Genomes Project reference panel (Phase 3, all ethnicities)^53^. Phasing was performed with the use of SHAPEIT (v2)^54^, and imputation was performed using minimac3^55^, minimac4, or IMPUTE (v2)^56^. Information on the study-specific genotyping, imputation, quality control, and analysis tools is provided in Supplementary Table 2. After genotype imputation, further quality control was applied to each study. SNPs with an imputation quality of r^2^ < 0.3 for minimac3 or minimac4, info < 0.4 for IMPUTE2 or an MAF of <0.01 were excluded.

### Association analysis of SNPs with daily alcohol intake and drinking status

Association analysis of SNPs with daily alcohol intake and drinking status was performed on three different subject groups: the entire population, subjects with the rs671 GG genotype only, and subjects with the rs671 GA genotype only. Because the number of carriers with the rs671 AA genotype was too small (Supplementary Table 3), association analysis in subjects with the rs671 AA genotype only was not conducted. Daily alcohol intake was base-2 log-transformed (log_2_ (grammes/day + 1)). The association of daily alcohol intake with SNP allele dose for each study was assessed by linear regression analysis with adjustment for age, age^2^, sex, and the first 10 principal components. The association of drinking status with SNP allele dose for each study was assessed by logistic regression analysis with adjustment for age, age^2^, sex, and the first 10 principal components. The effect sizes and standard errors estimated in the association analysis were used in the subsequent meta-analysis. The association analysis was conducted using EPACTS (http://genome.sph.umich.edu/wiki/EPACTS), SNPTEST^57^, or PLINK2^58^.

Association analysis, including interaction terms, was performed to evaluate the differential effects of each SNP on daily alcohol intake and drinking status between the GG and GA genotypes of rs671. In the interaction analysis for daily alcohol intake, the linear regression models were fit as:

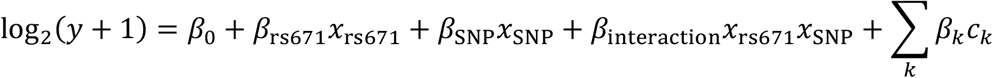

where *y* is daily alcohol intake (grammes/day). *x*_rs671_ is the genotype of rs671. The GG genotype is coded as 0, and the GA genotype is coded as 1. Carriers of the AA genotype were excluded from the analysis. *x*_SNP_ is the imputed genotype coded as [0,2] for each SNP. *c*_*k*_ is a covariate composed of age, age^2^, sex, and the first 10 principal components. The effect sizes of the interaction term, *β* _interaction_, and its standard errors estimated in the association analysis were used in the subsequent meta-analysis. In the interaction analysis for drinking status, the logistic regression model was fit as:

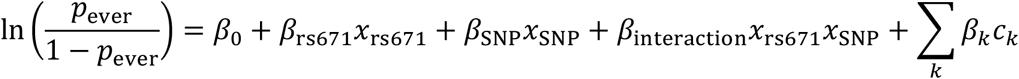

where *p*_ever_ is the probability that the subject is an ever drinker. Other variables and procedures are as above. The association analysis, including the interaction term, was conducted using PLINK2^58^. To identify studies with inflated GWAS significance, which can result from population stratification, we computed the genomic control λ ^59^. Before the meta-analysis, all study-specific results in the association analysis were corrected by multiplying the standard error of the effect size by λ if the λ of that study was greater than 1.

### Meta-analysis

The meta-analysis was performed with all Japanese subjects in the five cohorts (Supplementary Table 1). The results of association analyses for each SNP across the studies were combined with METAL software^60^ by the fixed-effects inverse-variance-weighted method. Heterogeneity of effect sizes was assessed by *I*^2^ and Cochran’s *Q* statistic. The meta-analysis included SNPs for which genotype data were available from at least three studies with a total sample size of at least 20,000 individuals for unstratified GWAS or interaction GWAS or 10,000 individuals for rs671-stratified GWAS. The genome-wide significance level α was set to a *P* value <5 × 10^−8^. *P*-values with <1.0×10^−300^ was calculated with Rmpfr of the R package.

### Functional annotations

To investigate the function of the lead SNP identified in this study, we adopted a series of bioinformatic approaches to collate functional annotations. We first used ANNOVAR^61^ to obtain an aggregate set of functional annotations — including gene locations and impacts of amino acid substitutions based on prediction tools, such as SIFT, PolyPhen-2, and CADD — for SNPs with *P* values <5 ×10^−8^. We also explored eQTLs in tissues considered relevant to daily alcohol intake and drinking status using the GTEx v8 database^62^ with regard to the loci identified in this study. The significant criteria for eQTL were based on the GTEx project: variants with a nominal *P* value below the gene-level threshold were regarded as significant. This threshold was determined by permutation tests in the GTEx project to keep the false discovery rate below 5%.

### Genotyping of rs56884502 and comparison with imputed genotype

*ALDH2* rs56884502 was further genotyped using TaqMan Assays on a 7500 Real-Time PCR System (Applied Biosystems, Foster City, CA, USA) in the selected 96 HERPACC samples which were also genotyped by Illumina HumanCoreExome. We confirmed a 100% match between the imputed and direct genotype data within these samples.

### Haplotype estimation of SNPs at 12q24

We estimated haplotypes from genotypes of rs56884502 and rs671 at 12q24 for the HERPACC (*n* = 96) and 1KGP-JPT^53^ (*n* = 104) samples. The genotype of rs56884502 for the HERPACC samples was determined by the method described above, while that of rs671 for these samples was determined by the Illumina HumanCoreExome SNP array. Furthermore, we estimated haplotypes from genotypes of rs671 and 73 SNPs at 12q24.12 which showed genome-wide significance for daily alcohol intake in the rs671 heterozygotes (GA) for the 1KGP-JPT samples (*n* = 104). Haplotype estimation was performed by the Haploview software^63^.

## Data Availability

Data will be available when the manuscript is formally published in certain journal.

## Acknowledgements

We are grateful to the participants and all staff members in this study. This study was supported by the National Cancer Center Research and Development Fund (28-A-19 and 31-A-18). The HERPACC Study was supported by Grants-in-Aid for Scientific Research from the Ministry of Education, Science, Sports, Culture and Technology of Japan Priority Areas of Cancer (17015018), Innovative Areas (221S0001), and JSPS KAKENHI Grants (JP16H06277 [CoBiA], JP26253041, JP20K10463) and Grant-in-Aid for the Third Term Comprehensive 10-year Strategy for Cancer Control from the Ministry of Health, Labour and Welfare of Japan. Yamagiwa-Yoshida Memorial UICC International Cancer Study Grants supported the initial component of the alcohol consumption GWAS within the HERPACC Study. The J-MICC Study was supported by Grants-in-Aid for Scientific Research for Priority Areas of Cancer (No. 17015018) and Innovative Areas (No. 221S0001) and by the Japan Society for the Promotion of Science (JSPS) KAKENHI Grant (No. 16H06277 [CoBiA]) from the Japanese Ministry of Education, Culture, Sports, Science and Technology. This work has also been supported in part by funding for the BioBank Japan Project from the Japan Agency for Medical Research and Development since April 2015, and the Ministry of Education, Culture, Sports, Science and Technology from April 2003 to March 2015. We thank Drs Nobuyuki Hamajima and Hideo Tanaka for their work in initiating and organizing the J-MICC Study as former principal investigators. The JPHC Study was supported by the National Cancer Center Research and Development Fund (23-A-31 [toku], 26-A-2, 28-A-19, 29-A-4 and 31-A-18), the Practical Research for Innovative Cancer Control (JP16ck0106095 and JP19ck0106266) from the Japan Agency for Medical Research and Development, and a Grant-in-Aid for Cancer Research from the Ministry of Health, Labour and Welfare of Japan (from 1989 to 2010). The TMM Study was supported by grants from the Reconstruction Agency, from the Ministry of Education, Culture, Sports, Science and Technology (MEXT), and from the Japan Agency for Medical Research and Development (AMED) [Grant no. JP20km0105001, JP20km0105002, JP20km0105003, and JP20km0105004]. The Nagahama Study was supported by JSPS, a Grant-in-Aid for Scientific Research (C), KAKENHI Grant Number JP17K07255 and JP17KT0125, and the Practical Research Project for Rare/Intractable Diseases from Japan Agency for Medical Research and Development, AMED, under Grant Numbers JP16ek0109070h0003, JP18kk0205008h0003, JP18kk0205001s0703 JP19ek0109283h0003, and JP19ek0109348h0002. The funders had no role in the design and conduct of the study; collection, management, analysis and interpretation of the data; preparation, review and approval of the article; or the decision to submit the article for publication. Where authors are identified as personnel of the International Agency for Research on Cancer / World Health Organization, the authors alone are responsible for the views expressed in this article and they do not necessarily represent the decisions, policy or views of the International Agency for Research on Cancer / World Health Organization.

## Ethical approval

All studies were approved by their respective institutional review boards.

## Conflicts of interest

The authors declare no potential conflicts of interest.

## Notes

### Competing Interest Statement

The authors have declared no competing interest.

### Funding Statement

This work was supported by the National Cancer Center Research and Development Fund (28-A-19 and 31-A-18). The HERPACC Study was supported by Grants-in-Aid for Scientific Research from the Ministry of Education, Science, Sports, Culture and Technology of Japan Priority Areas of Cancer (17015018), Innovative Areas (221S0001), and JSPS KAKENHI Grants (JP16H06277 [CoBiA], JP26253041, JP20K10463) and Grant-in-Aid for the Third Term Comprehensive 10-year Strategy for Cancer Control from the Ministry of Health, Labour and Welfare of Japan. Yamagiwa Yoshida Memorial UICC International Cancer Study Grants supported the initial component of the alcohol consumption GWAS within the HERPACC Study. The J-MICC Study was supported by Grants-in-Aid for Scientific Research for Priority Areas of Cancer (No. 17015018) and Innovative Areas (No. 221S0001) and by the Japan Society for the Promotion of Science (JSPS) KAKENHI Grant (No. 16H06277 [CoBiA]) from the Japanese Ministry of Education, Culture, Sports, Science and Technology. This work has also been supported in part by funding for the BioBank Japan Project from the Japan Agency for Medical Research and Development since April 2015, and the Ministry of Education, Culture, Sports, Science and Technology from April 2003 to March 2015. The JPHC Study was supported by the National Cancer Center Research and Development Fund (23-A-31 [toku], 26-A-2, 28-A-19, 29-A-4 and 31-A-18), the Practical Research for Innovative Cancer Control (JP16ck0106095 and JP19ck0106266) from the Japan Agency for Medical Research and Development, and a Grant-in-Aid for Cancer Research from the Ministry of Health, Labour and Welfare of Japan (from 1989 to 2010). The TMM Study was supported by grants from the Reconstruction Agency, from the Ministry of Education, Culture, Sports, Science and Technology (MEXT), and from the Japan Agency for Medical Research and Development (AMED) [Grant no. JP20km0105001, JP20km0105002, JP20km0105003, and JP20km0105004]. The Nagahama Study was supported by JSPS, a Grant-in-Aid for Scientific Research (C), KAKENHI Grant Number JP17K07255 and JP17KT0125, and the Practical Research Project for Rare/Intractable Diseases from Japan Agency for Medical Research and Development, AMED, under Grant Numbers JP16ek0109070h0003, JP18kk0205008h0003, JP18kk0205001s0703 JP19ek0109283h0003, and JP19ek0109348h0002.
The funders had no role in the design and conduct of the study; collection, management, analysis and interpretation of the data; preparation, review and approval of the article; or the decision to submit the article for publication.

### Author Declarations

The HERPACC Study was approved by the Institutional Ethics Committee of Aichi Cancer Center, Nagoya, Japan. The J-MICC Study was approved by the ethics committee of Nagoya University Graduate School of Medicine (Approval No.: 2010-0939), Nagoya, Japan. The JPHC Study was approved by the institutional review board of the National Cancer Centre (Approval No.:2011-044), Tokyo, Japan. The TMM CommCohort Study was approved by the Institutional Review Boards of Iwate Medical University and Tohoku University. The Nagahama Study was approved by the ethics committee of Kyoto University Graduate School of Medicine and by Nagahama Municipal Review Board.

