## Supplementary Information for "Genotype-stratified GWAS meta-analysis reveals novel loci associated with alcohol consumption"

Koyanagi, YN., Nakatochi, M. et al

**Supplementary Methods**

**Supplementary Tables** (Electronic Excel file)

Supplementary Table 1. Characteristics of study participants

Supplementary Table 2. Cohort-specific information on genotyping, imputation and association testing

Supplementary Table 3. Number of subjects with each genotype of rs671

Supplementary Table 4. Functional annotation results and allele frequencies across different ancestries for the lead SNPs

Supplementary Table 5. Haplotypes of rs56884502 and rs671 in Japanese samples

Supplementary Table 6. The 73 SNPs at 12q24.12 which showed genome-wide significance for daily alcohol intake in the rs671 heterozygotes (GA)

Supplementary Table 7. Associations of the previously reported loci in European populations other than the loci replicated with a genome-wide significance level in this study

Supplementary Table 8. Novel SNPs found to be eQTLs

**Details of studies**

Five studies with a total of 40,679 Japanese individuals with no history of cancer were included in the analysis of the present study. The individual study descriptions are as follows:

***Hospital-based Epidemiologic Research Program at Aichi Cancer Center (HERPACC) Study***

The HERPACC-2 Study was conducted between January 2001 and November 2005 at Aichi Cancer Center. Details of the HERPACC Study are described elsewhere[1]. Briefly, every first-visit outpatient ($n$ = 29,736) was asked to fill out a self-administered questionnaire regarding their lifestyle, including information on alcohol consumption, and provide a blood sample. 96.7% of eligible subjects ($n$ = 28,776) participated in the study with written informed consent and completed the questionnaire. 48.0% of participants ($n$ = 13,824) further provided 7mL of blood. Of these, 7,053 subjects were confirmed to have no detectable cancer and no history of neoplasia within a one-year window period from participation. For the present study, 4,958 randomly selected non-cancer subjects (genotyped using Illumina Human610-Quad [$n$ = 1,147] or Illumina HumanCoreExome [$n$ = 3,811]) were included in the final analysis, after quality control and exclusion of those who did not answer questions about alcohol consumption.

The HERPACC Study was conducted in accordance with the Declaration of Helsinki and approved by the Institutional Ethics Committee of Aichi Cancer Center, Nagoya, Japan.

***Japan Multi-Institutional Collaborative Cohort (J-MICC) Study***

The baseline survey of the J-MICC Study was conducted between 2004 and 2014 across 13 study sites in Japan. Details of the J-MICC Study are described elsewhere[2,3]. We selected 14,555 eligible participants for GWAS, considering the area distribution of respondents and the minimum sample size ($n$ = 500) for one study site. A total of 14,536 DNA samples were successfully genotyped for GWAS. After quality control, 14,091 participants remained for the imputation process. Participants who had a history of cancer or did not answer questions about alcohol drinking were excluded. Finally, 13,236 participants remained for the association analysis.

The J-MICC Study was approved by the ethics committee of Nagoya University Graduate School of Medicine (Approval No.: 2010-0939), Nagoya, Japan, and written informed consent was obtained from all the participants in the present study.

***Japan Public Health Center-based Prospective (JPHC) Study***

The JPHC Study was commenced in 1990 (cohort I) and 1993 (cohort II). Participants were recruited from 11 public health center areas in Japan. At the time of the baseline survey, the subjects were aged between 40-59 years in 1990 (cohort I) and 40-69 years in 1993 (cohort II). Details of the JPHC Study are described elsewhere[4]. From participants who answered the baseline survey questionnaire and donated 10mL blood in 9 public health center areas ($n$ = 33,736), we randomly selected 13,024 participants for genotyping from respective strata divided by sex, 5-year age category and public health center area. Subsequently, we performed the standard quality control for GWAS, leaving 10,295 participants for this analysis. After excluding participants who had a past history of cancer or did not completely answer questions about alcohol drinking, 10,037 participants (3,462 men and 6,575 women) were included in the analysis of drinking frequency and 9,142 participants (3,096 men and 6,046 women) were included in the analysis of drinking quantity.

Before the performance of genetic research on the JPHC samples, the study protocol was approved by the institutional review board of the National Cancer Centre (Approval No.:2011-044), Tokyo, Japan. Participants had the option to refuse participation in the research.

***The Tohoku Medical Megabank Community-Based Cohort (TMM CommCohort) Study***

The TMMCC Study[5] is a population-based cohort study which is part of the TMM Project[6], launched for the purpose of reconstruction from the Great East Japan Earthquake and establishment of personalized healthcare and medicine. The study recruited more than 80,000 residents aged 20 years or older living in Iwate and Miyagi Prefectures, located on the Pacific side of the Tohoku (northeastern) region of Honshu (the main island of Japan), from May 2013 to March 2016.

As a pre-analysis quality control, we filtered a data set of 9,965 participants (3,481 males and 6,484 females) who participated in the TMM CommCohort Study at health check-up venues in 2013, based on the following criteria: 1) call rate < 0.98, 2) inconsistent self-reported and genotype-based sex information, 3) close relatives (PI_HAT > 0.1875 = 3/16), 4) no available records for drinking-related items, height, or weight, 5) BMI ($kg/m^2$) < 14 or > 45, and 6) history of cancer. After filtering, 7,857 participants (2,691 males and 5,166 females) were retained for the current analysis.

The TMM CommCohort Study was approved by the Institutional Review Boards of Iwate Medical University and Tohoku University, and all the participants gave written informed consent. The study was conducted in accordance with the Declaration of Helsinki.

***The Nagahama Prospective Cohort for Comprehensive Human Bioscience (Nagahama) Study***

The Nagahama Study is a community-based cohort study conducted in Shiga, Japan[7]. Participants with no physical impairment or dysfunction were recruited from the general population aged 30 – 74 years in Nagahama City from 2008 to 2010. Of the 9,757 eligible participants, 5,730 randomly selected subjects who participated mainly during 2008 and 2009 were genotyped using illumine SNP array. We then obtained 4,591 subjects (Illumina Human610-Quad [*n* =1,170], Illumina Omni2.5 [*n* = 777], Illumina Omni5M [*n* = 472], Illumina Infinium HumanCoreExome [*n* =967] or Illumina Infinium Asian Screening Array [*n* =1,205]) by filtering with the following criteria: 1) call rate <0.99, 2) close relatives (PI_HAT > 0.38), and 3) history of cancer according to self-administrated questionnaire.

The Nagahama Study was approved by the ethics committee of Kyoto University Graduate School of Medicine and by Nagahama Municipal Review Board. All participants gave written informed consent.

**Phenotype**

Information on alcohol consumption was collected via a questionnaire in each study. Because the questionnaires were not homogeneous across the studies, we harmonized the two alcohol consumption phenotypes of drinking status (never versus ever drinker) and daily alcohol intake (g/day) in accordance with each study's criterion, as follows:

***The HERPACC Study***

In the questionnaire related to alcohol consumption in the HERPACC-2 Study, the participants were first asked about drinking status, as "never" (including "almost never"), "former", or "current". Participants who selected "former" or "current" were defined as ever drinkers. Former and current drinkers were then asked about the frequency of alcohol drinking (<1 day/week, 1–2 days/week, 3–4 days/week, or ≥5 days/week), along with type of beverage (Japanese sake, beer, shochu, whiskey, and wine) and average consumption during each drinking session based on their average drinking behavior. To calculate daily alcohol intake (g/day), each category of drinking frequency was assigned a score as follows: 0 for never drinkers, 0.5 for <1 day/week, 1.5 for 1–2 days/week, 3.5 for 3–4 days/week, and 6 for ≥5 days/week. Alcohol intake per drinking session was estimated based on the concentration of ethanol in each beverage. In the HERPACC Study, one unit of drink was assumed to contain 23 g of ethanol for 180 ml (one "go") of Japanese sake, 633 ml (one large bottle) of beer, 90 ml (a half "go") of shochu (distilled spirit), 60 ml (double shot) of whiskey, and 200 ml (two and a half glasses) of wine. We therefore estimated alcohol intake per drinking session by multiplying the summed units of each type of beverage by 23. Finally, daily alcohol intake was estimated by multiplying alcohol intake per drinking session by the frequency score/7.

***The J-MICC Study***

All of the study sites used a common, standard self-administered questionnaire. The participants were first asked about drinking status (almost none, former, or current). Those who selected "almost none" were defined as never drinkers and the others were defined as ever drinkers. Current drinkers were further required to report the frequency (almost none, 1 to 3 days/month, 1 to 2 days/week, 3 to 4 days/week, 5 to 6 days/week, or everyday) and amount of alcohol consumption for six alcoholic beverages (Japanese sake, shochu, shochu-based highball, beer, whisky, and wine). Daily alcohol intake was estimated for current drinkers based on the frequencies and amount of each type of alcoholic beverage consumed over the past year.

***The JPHC Study***

Information on alcohol intake was obtained regarding frequency and quantity using a validated questionnaire at baseline survey. The average frequency of alcohol intake was reported according to six categories for cohort I: "almost never", "1–3 days per month", "1–2 days per week", "3–4 days per week", "5–6 days per week", and "every day". Participants who selected "almost never" were defined as never drinkers and others were defined as ever drinkers. Type of alcohol and the average amount were also ascertained. Participants in cohort II were asked to indicate their alcohol drinking status as "never", "past", or "current drinker". Past and current drinkers provided information on the average frequency of intake, the type of alcohol consumed, and the average quantity consumed per day. To calculate the total alcohol consumption per day, we added the quantity of ethanol of each alcoholic beverage. The amount of ethanol was assigned as follows: 180 ml of sake (rice wine) was regarded as 23 g of ethanol, 180 ml of shochu or awamori (white spirits) as 36 g, 633 ml of beer as 23 g, 30 ml of whiskey or brandy as 10 g, and 60 ml of wine as 6 g. This calculation was validated in the previous paper[8].

***The TMMCC Study***

Drinking status for each participant was assigned based on a question on drinking habit in a self-administered questionnaire used in the cohort. The question has four options: "current drinker", "former drinker", "(almost) non-drinker", and "alcohol intolerance". Both "(almost) non-drinker" and "alcohol intolerance" were classified as "never drinker"; the others were classified as "ever drinker". The participants were also asked to answer a pair of questions, one on drinking frequency and the other on amount per event, for each type of alcoholic beverage. Alcohol intake was calculated for 7,627 participants (3,634 current drinkers and 3,993 never-drinkers), with 230 former drinkers excluded. The question on drinking frequency had six options: "almost none", "1-3 days/month", "1-2 days/week", "3-4 days/week", "5-6 days/week", and "every day." The answers were converted into the number of events per day, i.e., 0, 2/30, 1.5/7, 1/2, 5.5/7, and 1, respectively. The amount per event was converted into the ethanol content (in grams) as follows: 180 ml of sake (rice wine) as 23 g, 180 ml of shochu (distilled spirits) as 36 g, 180 ml of chuhai (shochu-based beverage) as 12.96 g, 633 ml of beer as 23 g, 30 ml of whisky as 10 g, and 100 ml of wine as 12 g. The alcohol intake (g/day) for each participant was then calculated as the sum of the product of the ethanol content and the number of events per day for all types of alcoholic beverage. For never drinkers, alcohol intake was considered as zero.

***The Nagahama Study***

Information on the amount and frequency of alcohol consumption was collected through questionnaires. Participants were first asked to report their drinking status (never, almost never, former, sometimes, or every day). The participants who selected "never" or "almost never" were defined as never drinkers and the others were defined as ever drinkers. Those who selected "sometimes" or "everyday" were regarded as "current" drinkers, and were further required to report the frequency and amount of alcohol consumption for six alcoholic beverages (Japanese sake, shochu, shochu-based highball, beer, whisky, and wine). Daily alcohol intake was estimated for current drinkers based on the consumption frequency and amount of each type of alcoholic beverage over the past year.

**Supplementary Figure 1. Q-Q plots for rs671 genotype-stratified GWAS meta-analysis of daily alcohol intake**

The results for (a) rs671 wild-type homozygotes (GG), (b) rs671 heterozygotes (GA), (c) unstratified, and (d) interaction with rs671 are shown. The vertical and horizontal axes indicate the observed and expected $-\log_{10}(P \text{ value})$ for tests of association between SNPs and daily alcohol intake, respectively.

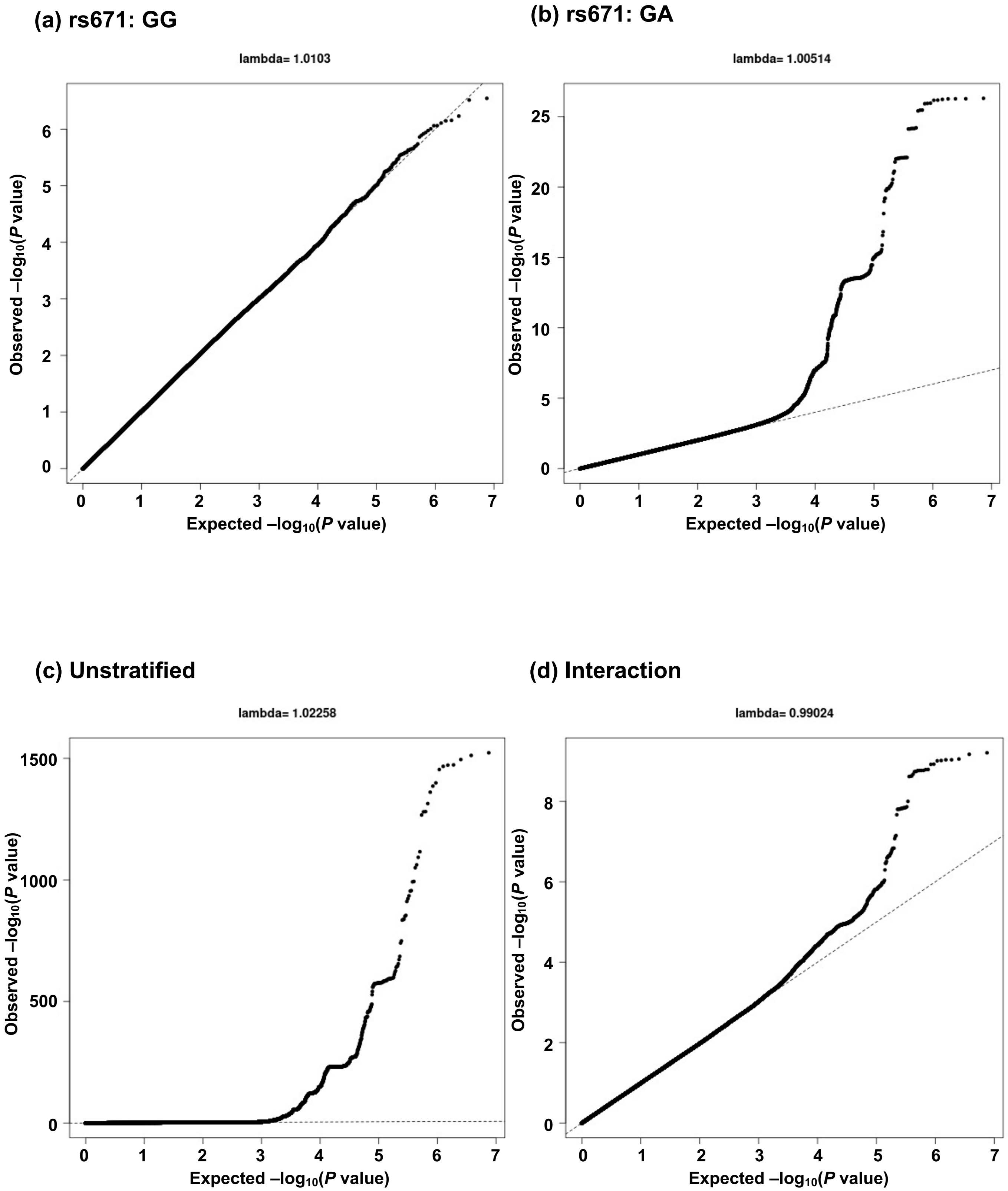

**Supplementary Figure 2. Q-Q plots for rs671 genotype-stratified GWAS meta-analysis of drinking status**

The results for (a) rs671 wild-type homozygotes (GG), (b) rs671 heterozygotes (GA), (c) unstratified, and (d) interaction with rs671 are shown. The vertical and horizontal axes indicate the observed and expected –$\log_{10}$($P$ value) for tests of association between SNPs and drinking status, respectively.

**(a) rs671: GG**

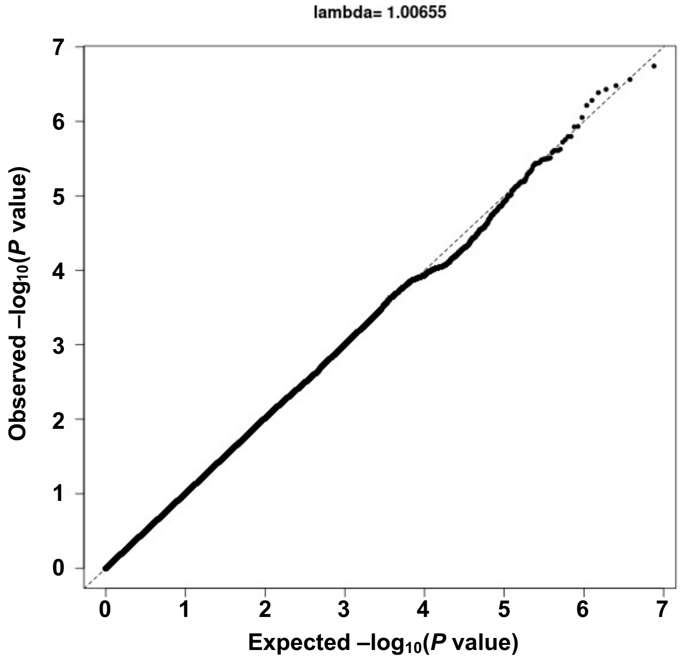

**(b) rs671: GA**

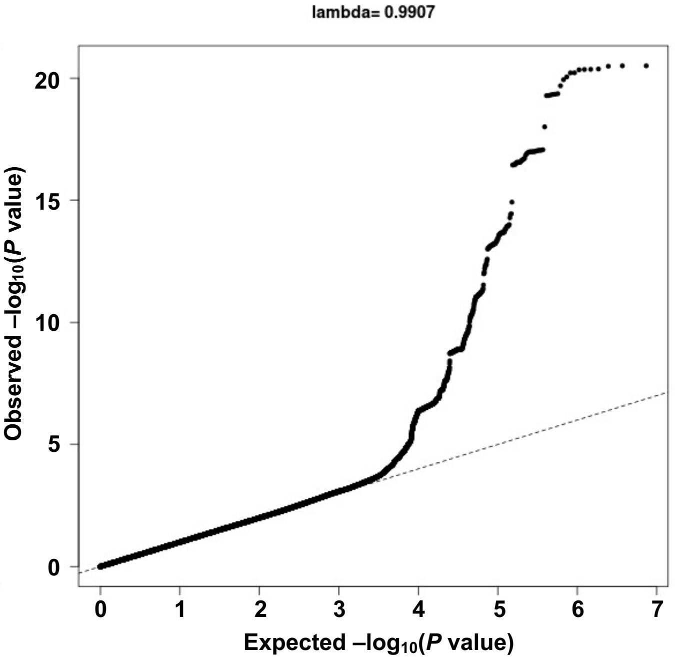

**(c) Unstratified**

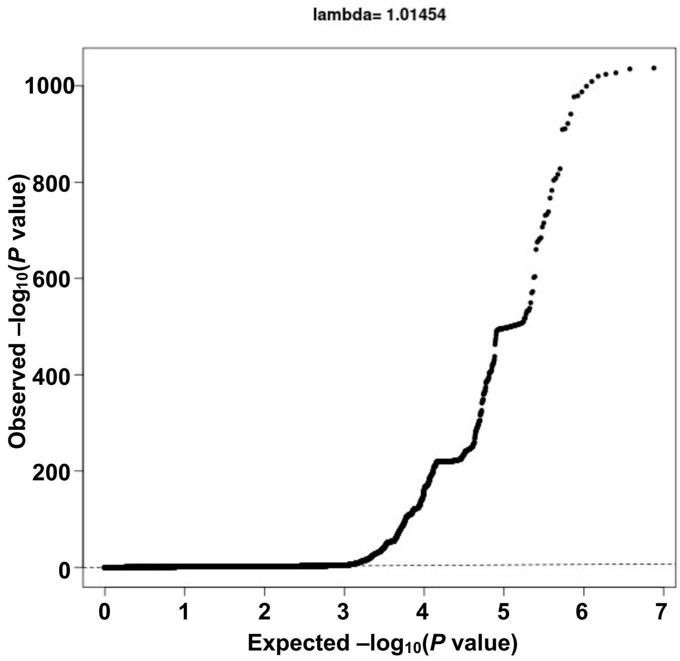

**(d) Interaction**

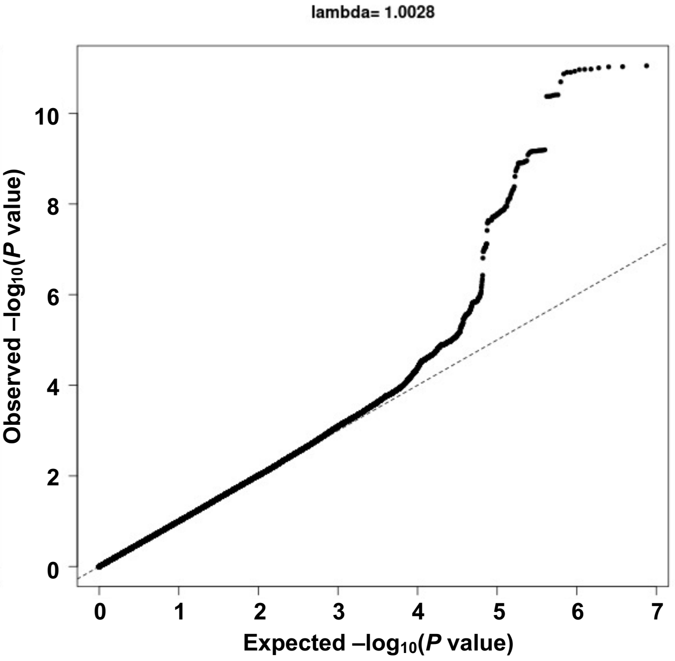

**Supplementary Figure 3. Regional association plots of the identified regions for daily alcohol intake other than those shown in Figure 3**

Regional association plots for (a) rs671 heterozygotes (GA), (b) unstratified, and (c) interaction with rs671 are shown. The vertical axis indicates the $-\log_{10}(P\text{ value})$ for assessment of the association of each SNP with daily alcohol intake or drinking status. The black line represents a genome-wide significance threshold of $5.0\times10^{-8}$. The colors indicate the LD ($r^2$) between each lead SNP and neighboring SNPs based on the JPT population in the 1000 Genomes Project Phase 3.

## (a) rs671:GA

### 4q23 (rs10005290)

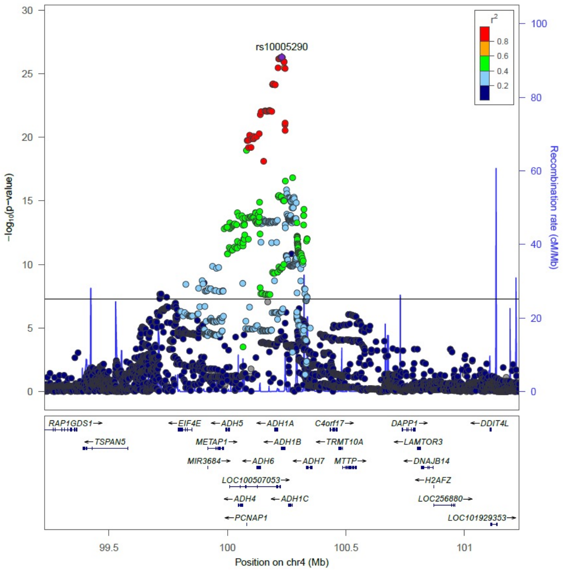

### 9p13.2 (rs2228093)

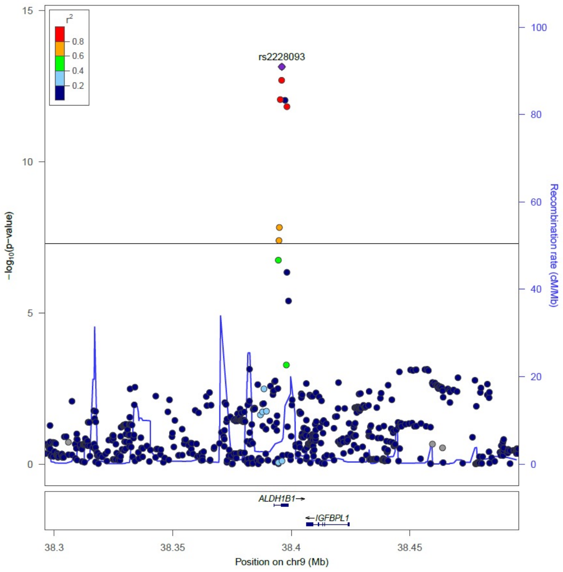

### 9q21.13 (rs8187929)

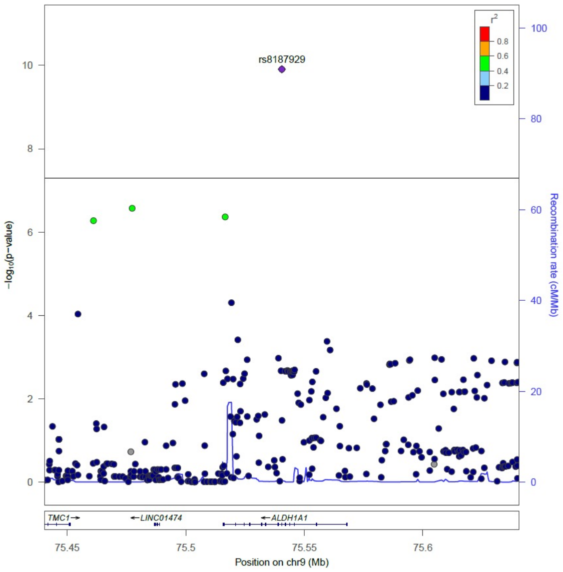

### (b) Unstratified

### 2p23.3 (rs1260326)

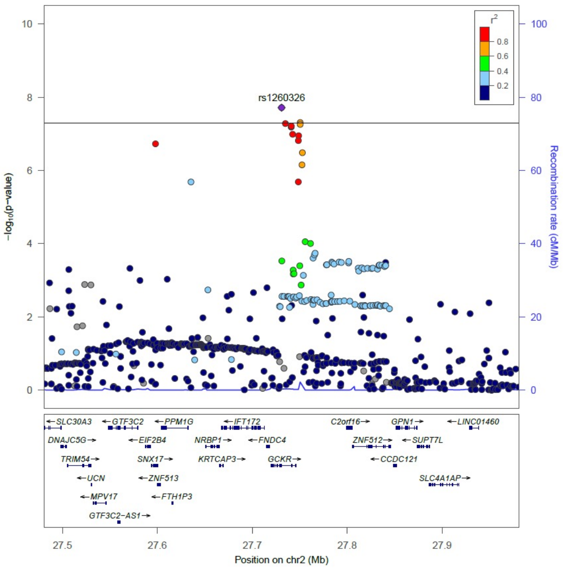

### 4q23 (rs1229984)

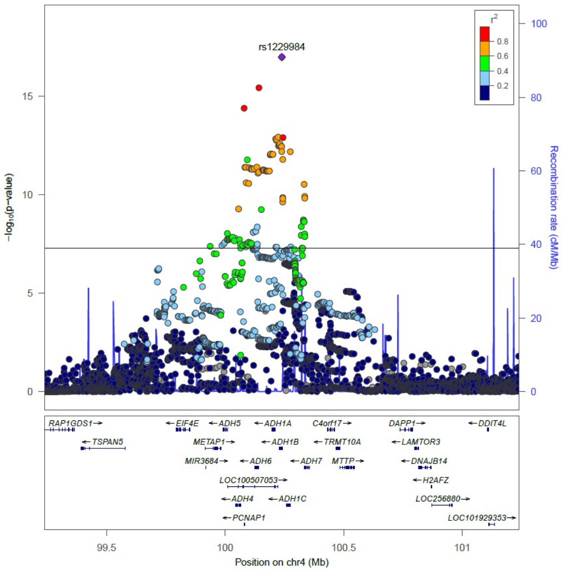

### 12q24.12 (rs671)

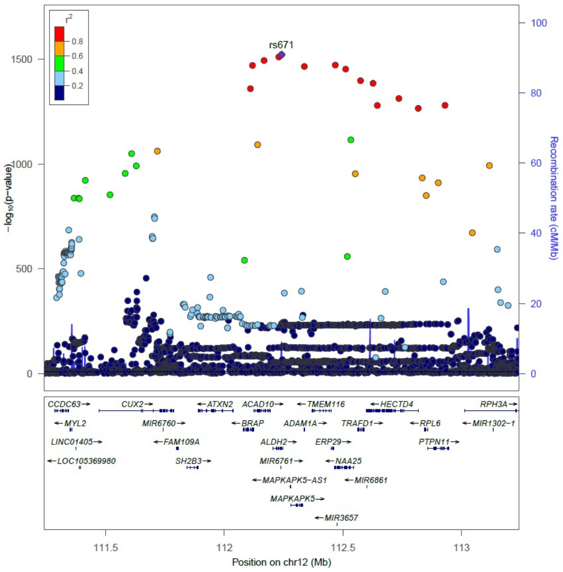

### (c) Interaction

### 4q23 (rs1813977)

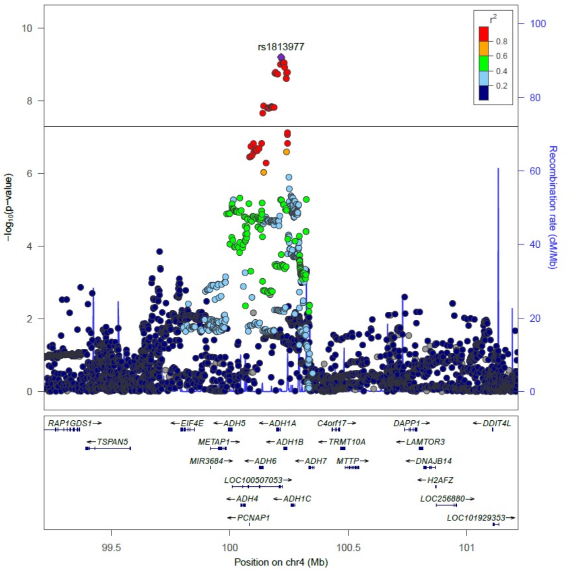

### 9p13.2 (rs2228093)

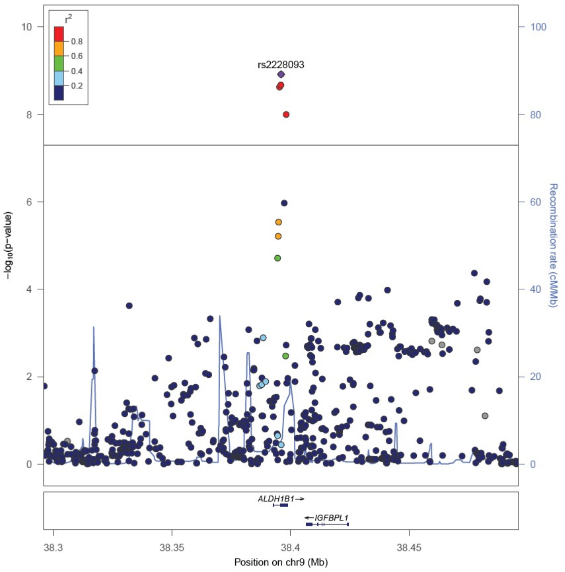

**Supplementary Figure 4. Regional association plots of the identified regions for drinking status other than those shown in Figure 3**

Regional association plots for (a) rs671 heterozygotes (GA), (b) unstratified, and (c) interaction with rs671 are shown. The vertical axis indicates the $-\log_{10}(P\text{ value})$ for the assessment of the association of each SNP with daily alcohol intake or drinking status. The black line represents a genome-wide significance threshold of $5.0\times10^{-8}$. The colors indicate the LD ($r^2$) between each lead SNP and neighboring SNPs based on the JPT population in the 1000 Genomes Project Phase 3.

## (a) rs671:GA

### 4q23 (rs35333426)

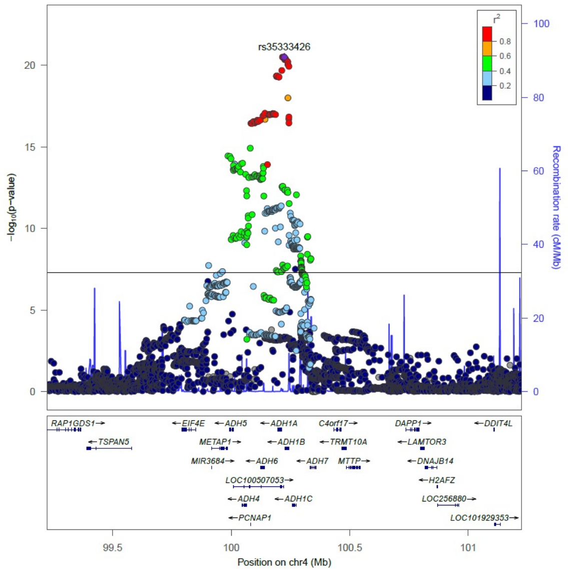

### 9p13.2 (rs2228093)

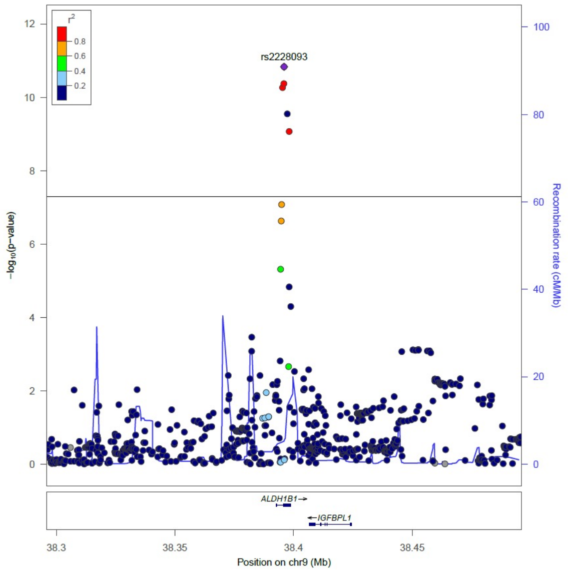

### 9q21.13 (rs8187929)

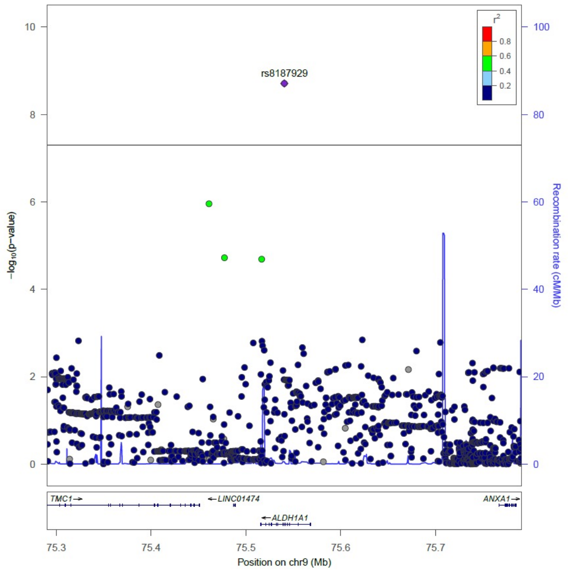

### (b) Unstratified

### 4q23 (rs1229984)

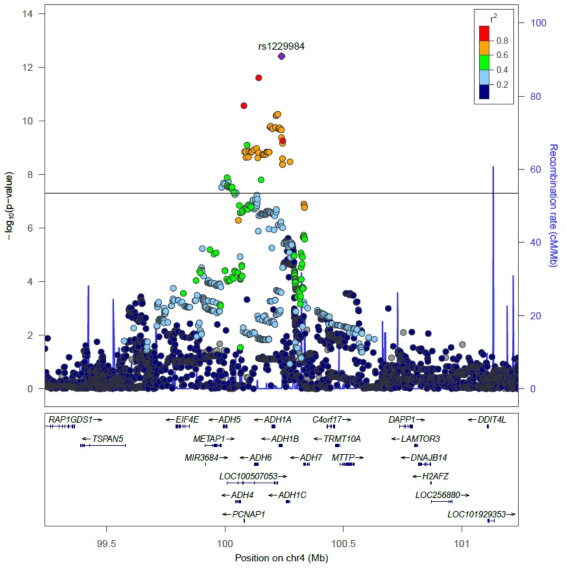

### 12q24.12 (rs671)

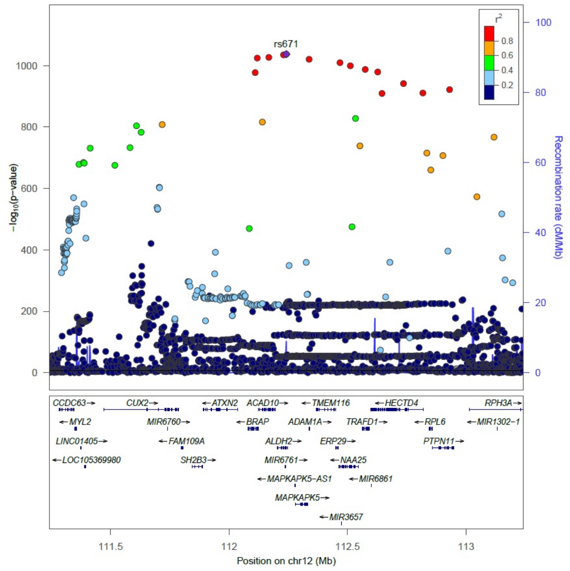

### (c) Interaction

### 4q23 (rs10005290)

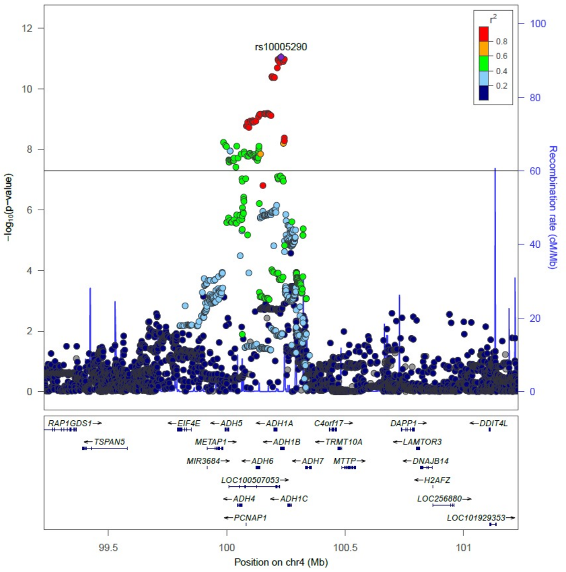

### 9p13.2 (rs2228093)

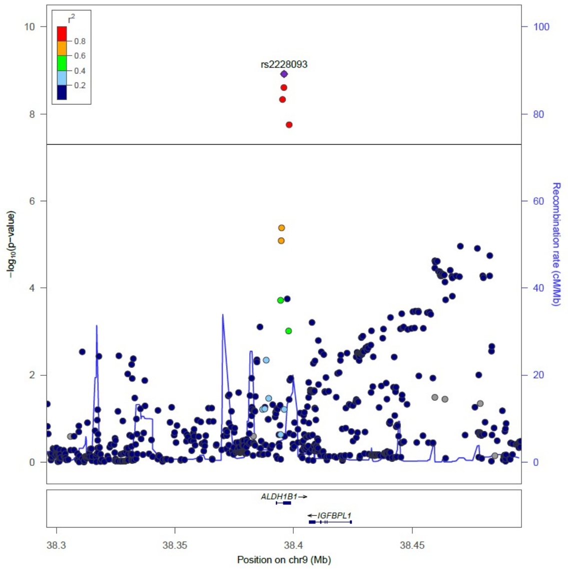

**Supplementary Figure 5. LD maps of rs671 and 73 SNPs with genome-wide significance with daily alcohol intake for rs671 heterozygotes (GA) at 12q24 based on the 1000 Genomes Phase 3 JPT**

Pairwise linkage disequilibrium (a) $r^2$ values (white to black scales indicate low to high values) and (b) *D'* values (white to red scales indicate low to high values), as determined using Haploview software. The SNPs surrounded by red rectangles indicate rs7978737, rs56884502, and rs671.

**(a) $r^2$**

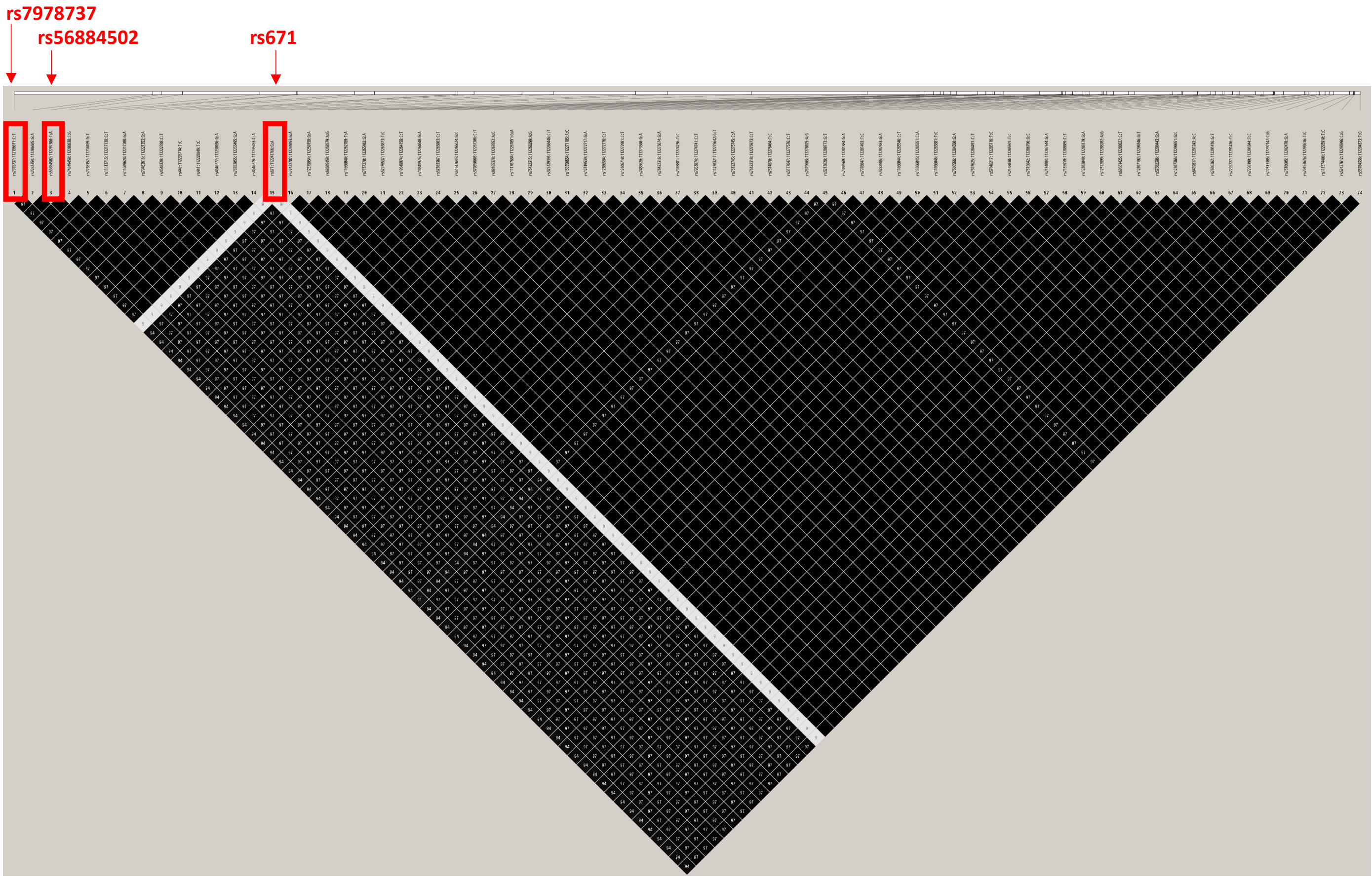

**(b) *D’***

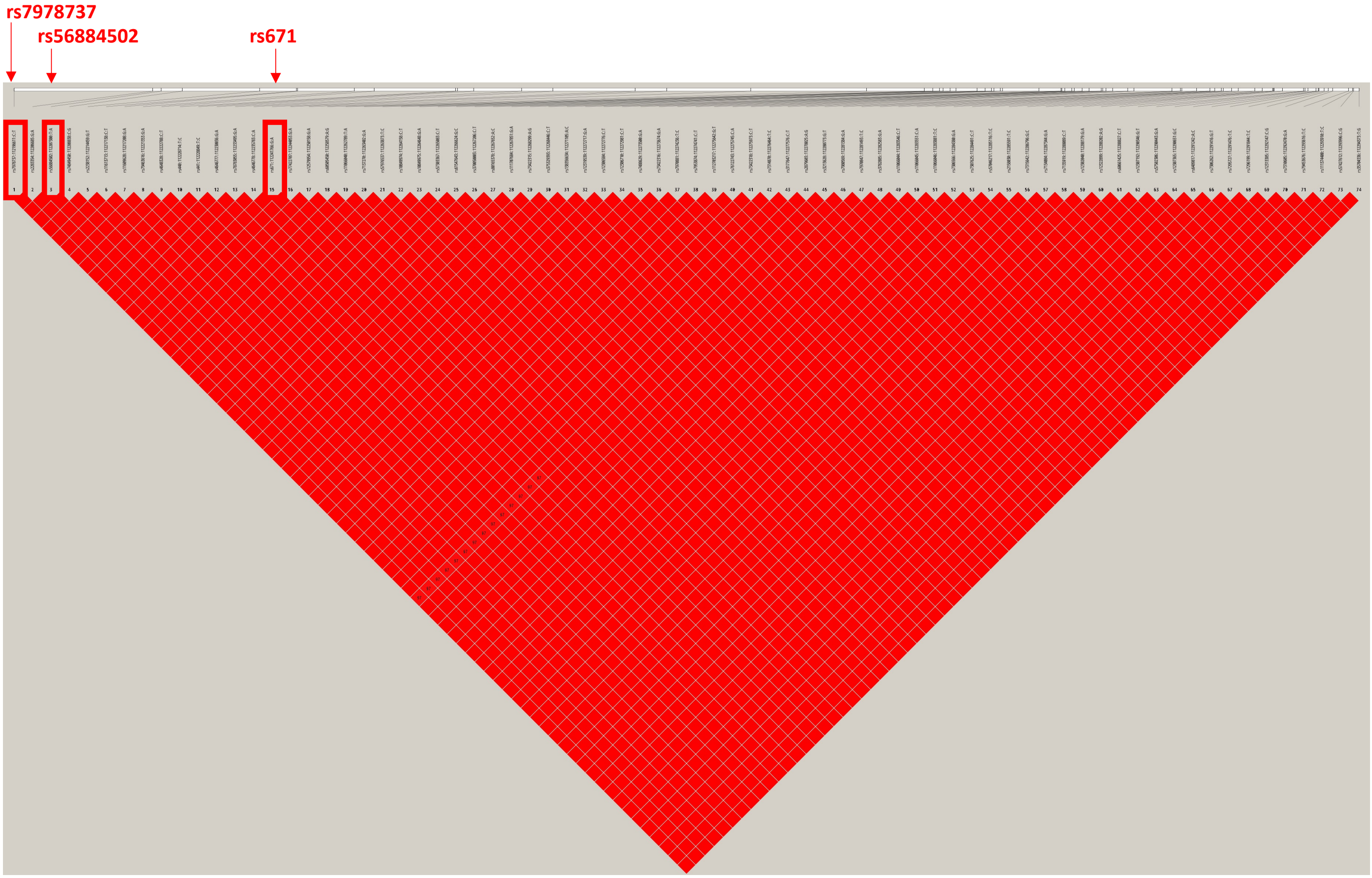

**Supplementary Figure 6. Haplotypes of rs671 and 73 SNPs at 12q24 with genome-wide significance with daily alcohol intake for rs671 heterozygotes (GA) based on the 1000 Genomes Phase 3 JPT**

The upper numbers correspond to the SNP numbers shown in Supplementary Figure 5. The haplotype surrounded by the blue rectangle indicates that with the A allele of rs671. The haplotypes were estimated using the Haploview software. These three haplotypes explain >99%.

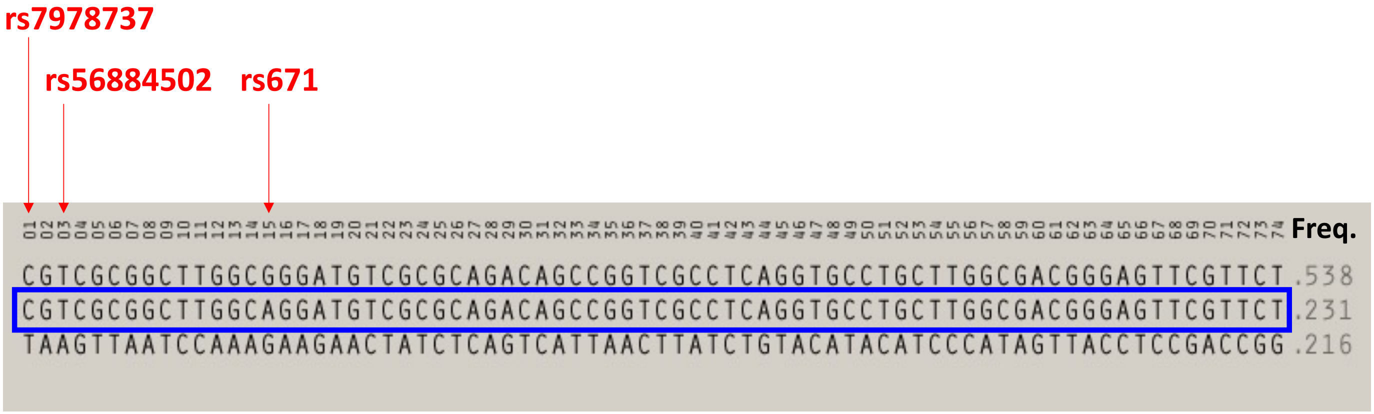
